# Combining AI and human support in mental health: a digital intervention with comparable effectiveness to human-delivered care

**DOI:** 10.1101/2024.07.17.24310551

**Authors:** Clare E Palmer, Emily Marshall, Edward Millgate, Graham Warren, Michael P. Ewbank, Elisa Cooper, Samantha Lawes, Malika Bouazzaoui, Alastair Smith, Chris Hutchins-Joss, Jessica Young, Morad Margoum, Sandra Healey, Louise Marshall, Shaun Mehew, Ronan Cummins, Valentin Tablan, Ana Catarino, Andrew E Welchman, Andrew D Blackwell

## Abstract

**Background:** Escalating global mental health demand exceeds existing clinical capacity. Scalable digital solutions will be essential to expand access to high-quality mental healthcare for everyone. This study evaluated a structured, evidence-based digital program for mild, moderate and severe anxiety that combined an Artificial Intelligence (AI) driven conversational agent to deliver content with human clinical oversight and user support to maximize outcomes.

**Objective:** This study aimed to measure engagement, clinical effectiveness, acceptability and safety of this digital intervention in comparison to externally generated comparator groups.

**Methods:** All prospective participants (N=299) were given the digital intervention to use for up to 9 weeks. Endpoints for effectiveness, engagement, acceptability, and safety were collected before, during and after the intervention, and at one-month follow-up. Adherence and effectiveness were compared to three propensity-matched real-world patient comparator groups: i) waiting control; ii) face-to-face cognitive behavioral therapy (CBT); and iii) remote typed-CBT.

**Results:** Participants used the program for a median of 6 hours over 53 days. There was a large clinically meaningful reduction in anxiety symptoms for the intervention group (per-protocol (PP; n=169): change on GAD-7 = –7.4, *d* = 1.6; intention-to-treat (ITT; n=299): change on GAD-7 = –5.4, *d* = 1.1) that was statistically superior to the waiting control (PP: *d* = 1.3; ITT: *d* = 0.8), non-inferior to human-delivered care, and was sustained at one-month follow-up.

**Conclusions:** By combining AI and human support, the digital intervention achieved clinical outcomes comparable to human-delivered care while significantly reducing the required clinician time by up to 8 times. These findings highlight the potential of technology to scale effective evidence-based mental healthcare, address unmet need, and ultimately impact quality of life and economic burden globally.

## Introduction

Mental health conditions are one of the economic and healthcare challenges of our time. Globally, one in eight people live with a mental health condition^1^, yet only one in four who require treatment receive it^2^. Advances in technology and widespread internet access have been pivotal in increasing access to high-quality mental healthcare. However, one-to-one mental healthcare is inherently limited in its ability to meet the rising mental health demand, and there remains a significant shortage of therapists: there are only four psychiatrists per 100,000 people globally^3^, and 58% of the US population live within a health workforce shortage area^4^. Technology is primed to enable massive scaling of mental health interventions to increase both access and quality of support worldwide^5^.

Rapid advances in computing and Artificial Intelligence (AI) in recent years have led to a rise in the development of digital interventions aiming to solve this scalability problem, and there are an estimated 10,000–20,000 smartphone applications available for mental health support^6,7^. These solutions have the potential to enable timely access to support when needed, negate the logistical challenges of attending regular appointments, offer greater patient choice, and reduce burden on therapists and healthcare services^8^. However, real-world usage of many digital mental health solutions – most of which are self-led - has been poor^9–11^. Despite a reported willingness of patients to adopt smartphone applications^12^, one month retention rates are typically under 6%^13^. Moreover, a recent meta-analysis of mental health applications for symptoms of anxiety and depression found a small pooled clinical effect size (*g*=0.26) and highlighted that only 48% delivered content based on Cognitive Behavioral Therapy (CBT) principles^14^ – a “gold-standard” evidence-based approach for anxiety and depression^15^. Improving access is crucial, but equally vital is ensuring the support available to patients is engaging and effective.

NHS Talking Therapies (NHS TT, formerly IAPT) is a world-leading initiative designed to increase access to and improve delivery of mental health treatment in the UK. Fundamental to the success of NHS TT is systematic outcomes-monitoring, use of evidence-based treatment protocols, and an appropriately trained and supervised workforce^16^. The acceleration of telehealth and expansion of care delivery through digital platforms (e.g. typed conversations) has also enabled insights into the relationship between the active components of evidence-based treatments and clinical outcomes^17,18^. Combining this approach with the scalable, systematic delivery of evidence-based protocols through digital tools, offers the opportunity to reduce heterogeneity across the provision of mental healthcare worldwide, and accelerate large-scale scientific research to further enhance treatment quality and personalization^19^. High-quality, accessible digital mental healthcare has the potential to maximize impact globally by both improving patient quality of life and reducing the growing economic burden of mental health on health systems and society^20,21^.

In this study, we evaluated a digital program that uses this approach to alleviate mild, moderate and severe symptoms of generalized anxiety in adults. The program was designed to maximize engagement and effectiveness by using i) a structured evidence-based program drawing on principles from traditional CBT^22^ including third wave approaches i.e. Acceptance and Commitment Therapy (ACT)^23^, and ii) an AI-powered conversational agent to deliver the program content in a personalized way. In addition, a dedicated human clinical and user support service was designed to wrap around the digital program, following previous research that human support significantly improves engagement with digital interventions^12,24^. This service was developed to provide appropriate support while maintaining the scalability of the digital solution.

This study aimed to measure engagement, clinical effectiveness, acceptability and safety of this digital intervention. Evidence of the effectiveness of a digital intervention is often established through the comparison between the intervention and a waitlist control or self-led non-digital treatment only. However, if digital programs are to provide a scalable solution to global mental health need, we should expect them to provide comparable effectiveness to current standards of care. In this pragmatic, prospective single-intervention arm study, we compared the digital program against propensity-matched external control data from three groups of real-world NHS patients: i) a waiting control with no intervention; ii) patients receiving human-delivered face-to-face CBT; and iii) patients receiving human-delivered typed-CBT. While 1:1 face-to-face therapy serves as the gold-standard for comparison, 1:1 typed-therapy provides a more analogous comparison to the digital program under evaluation where content is predominantly delivered through written communication with the conversational agent. This study design allowed us to evaluate the comparative clinical effectiveness of the digital intervention to human-delivered standard care.

## Methods

### Study design

This was a pragmatic, single-intervention arm study with multiple external control groups to measure the engagement, clinical effectiveness, acceptability and safety of a digital program to alleviate symptoms of generalized anxiety in a sample of 300 UK participants. This study was conducted by ieso Digital Health (“ieso”, https://www.iesogroup.com/), an outpatient service provider within NHS TT delivering 1:1 human-delivered CBT via a typed modality to treat patients with common mental health disorders. The digital program evaluated here (software name: IDH-DP2-001) was developed by ieso as part of a clinical innovation program creating new scalable digital solutions for mental health support. This was an externally controlled trial meaning comparator arms (sometimes referred to as synthetic control arms) were generated through 1:1 propensity-matching of participants with real-world patients. External propensity-matched control groups were generated to evaluate the digital intervention in comparison to no intervention (i.e. waiting control), face-to-face CBT (gold-standard benchmark), and typed-CBT. This latter group provides an important comparator as it is an example of human-delivered care that closely mirrors the written content delivery within the digital program.

The intervention was delivered via a smartphone application (iPhone & Android). Following an initial clinical assessment with a qualified clinician, eligible participants downloaded the software on their personal smartphone and completed the program in their own time and according to a defined schedule. Participants were required to complete the six-module program within nine weeks.

At the point of consent, all participants were asked if they were willing to participate in interviews with additional compensation offered. The sub-sample (based on first-come-first-served sign-up for available interview slots) attended a semi-structured interview pre- and post-intervention to gather qualitative insights into the experience, acceptability and perceived safety of the digital program. Findings on acceptability of the digital program from detailed qualitative analysis of these interviews are reported in a separate publication.

The study was pre-registered (ISRCTN ID: 52546704) and obtained ethical approval prior to recruitment (IRAS ID: 327897, NHS Research Ethics Committee: West of Scotland REC 4). The trial design and participant CONSORT flowchart^25^ are summarized in Figure 1. In line with the Declaration of Helsinki, all participants provided signed informed consent and were debriefed following the study. This study was conducted in accordance with Good Clinical Practice (GCP) principles.

**Figure 1.**
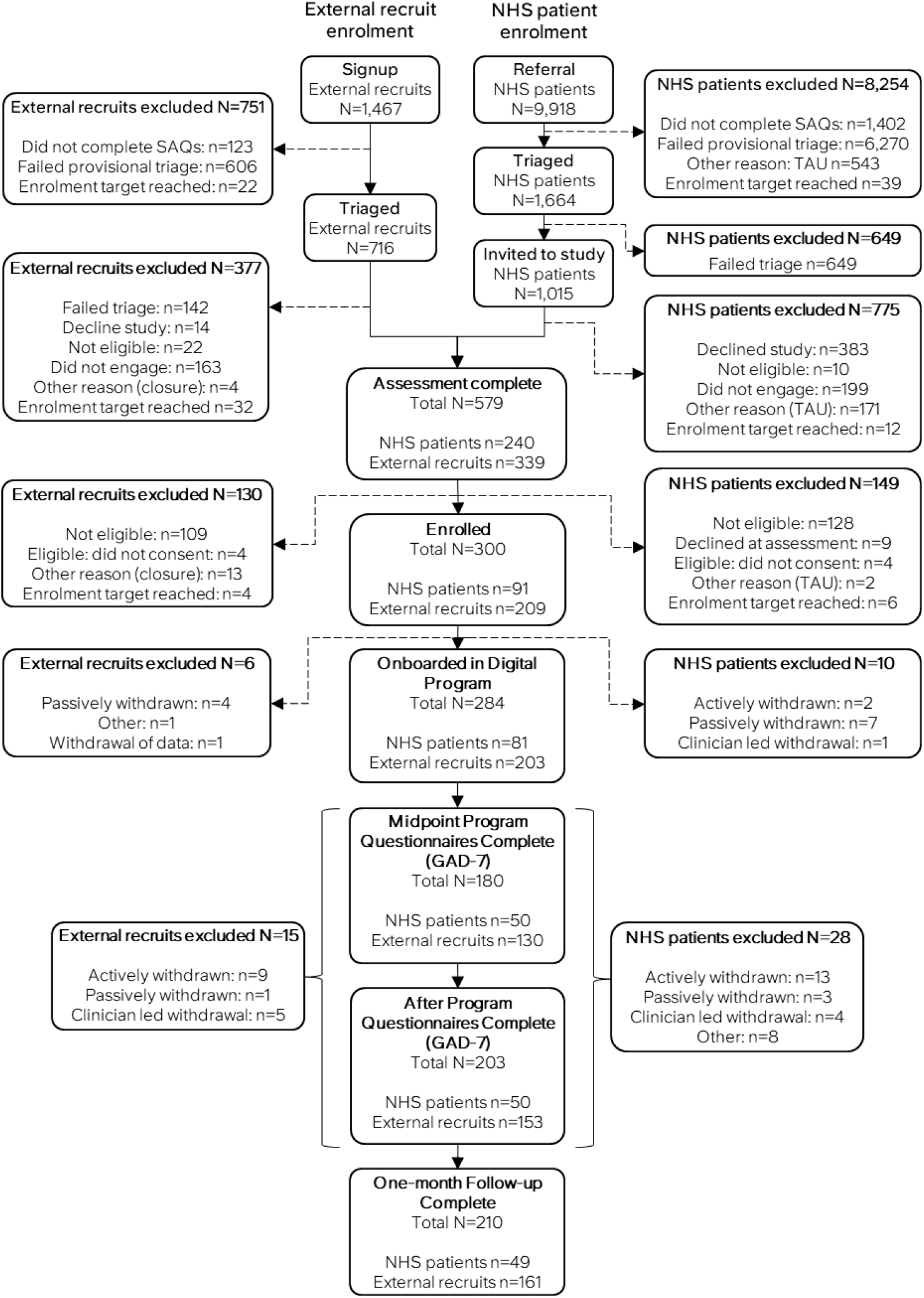
CONSORT diagram. Prior to assessment enrolment avenues differed for external recruits (left) and patients referred to ieso for typed therapy (either from an NHS Provider or via a self-direct referral; right). External recruits signed-up specifically for the study via an external webpage following social media or email advertisements. All potential participants irrespective of enrolment avenue were triaged for suitability based on a Self-Assessment Questionnaire (SAQ). For patients, only those deemed to be potentially eligible were invited to participate. Participants were withdrawn either actively (requested to withdraw), passively (dropped-out or disengaged from study procedures), clinician-led (withdrawn based on clinician recommendation), or other (due to reasons such as technical issues). TAU = treatment as usual.

### Study endpoints & data capture

Anxiety and mood symptoms were measured before and after the intervention, as well as at the beginning of each module within the program (maximum 6 symptom checkins) using the Generalized Anxiety Disorder-7 scale (GAD-7)^26^ and the Patient Health Questionnaire (PHQ-9)^27^ scale. The Work and Social Adjustment Scale (WSAS)^28^ and the inflexibility scale (30 items) of the Multidimensional Psychological Flexibility Inventory (MPFI)^29^ were collected pre-intervention, at the program mid-point and post-intervention, as measures of functioning and psychological inflexibility, respectively. The following validated self-report measures were collected only at post intervention: the User Engagement Scale (UES)^30^, the System Usability Scale (SUS)^31^, and the Service-User Technology Acceptability Questionnaire (SUTAQ)^32^. A qualitative feedback survey was also administered post-intervention and at one-month follow-up. Demographic data were collected at enrolment and are summarized in Table 1. Findings from the SUS, UES, SUTAQ, MPFI, feedback surveys and qualitative data from pre- and post-intervention semi-structured interviews are reported in a separate publication. Safety endpoints were serious adverse events, software deficiencies, and number of cases withdrawn based on clinician assessment of suitability to continue with the program. Software deficiencies included malfunctions or errors of the software that could result in issues related to safety or software performance. Serious adverse events were defined as any adverse event that led to death or serious deterioration in a participant’s health.

**Table 1.** Sample characteristics of the digital intervention group for both ITT and PP samples.

| Demographic | Category | ITT<br>(N=299) | PP<br>(N=169) |
| --- | --- | --- | --- |
| Age, Mean (SD) | - | 39.8 (12.8) | 41.7 (11.8) |
| Baseline GAD-7, Mean (SD) | - | 12.5 (3.3) | 12.4 (3.4) |
| Baseline PHQ-9, Mean (SD) | - | 8.0 (3.7) | 8.0 (3.8) |
| Gender, n (%) | Female | 240 (80.3) | 137 (81.1) |
|  | Male | 46 (15.4) | 26 (15.4) |
|  | Other | 4 (1.3) | 2 (1.2) |
|  | Not Known | 9 (3.0) | 4 (2.4) |
| Ethnicity, n (%) | White | 266 (89.0) | 155 (91.7) |
|  | Mixed | 5 (1.7) | 2 (1.2) |
|  | Asian | 14 (4.7) | 6 (3.6) |
|  | Black/African/Caribbean/Black British | 3 (1.0) | 1 (0.6) |
|  | Other | 2 (0.7) | 1 (0.6) |
|  | Prefer not to say | 9 (3.0) | 4 (2.4) |
| Highest Qualification, n (%) | Post-graduate degree level qualification | 103 (34.4) | 65 (38.5) |
|  | Degree level qualification | 100 (33.4) | 59 (34.9) |
|  | Qualifications below degree level | 84 (28.1) | 41 (24.3) |
|  | No formal qualifications | 2 (0.7) | 1 (0.6) |
|  | Don't know | 7 (2.3) | 2 (1.2) |
|  | Other | 1 (0.3) | 0 (0) |
|  | Prefer not to say | 2 (0.7) | 1 (0.6) |
| Disability, n (%) | Disability | 56 (18.7) | 33 (19.5) |
|  | No Perceived Disability | 232 (77.6) | 132 (78.1) |
|  | Prefer not to say | 11 (3.7) | 4 (2.4) |
| Chronic health condition, n (%) | Yes | 114 (38.1) | 70 (41.4) |
|  | No | 167 (55.9) | 91 (53.8) |
|  | Not Known | 18 (6.0) | 8 (4.7) |
| Religion, n (%) | No religion | 187 (62.5) | 104 (61.5) |
|  | Christian | 71 (23.7) | 45 (26.6) |
|  | Buddhist | 1 (0.3) | 1 (0.6) |
|  | Hindu | 5 (1.7) | 3 (1.8) |
|  | Jewish | 3 (1.0) | 1 (0.6) |
|  | Muslim | 5 (1.7) | 0 (0) |
|  | Sikh | 1 (0.3) | 1 (0.6) |
|  | Other | 11 (3.7) | 7 (4.1) |
|  | Prefer not to say | 15 (5.0) | 7 (4.1) |
| Sexual Orientation, n (%) | Heterosexual | 237 (79.3) | 132 (78.1) |
|  | Gay/Lesbian | 7 (2.3) | 5 (3.0) |
|  | Bi-sexual | 32 (10.7) | 22 (13.0) |
|  | Other sexual orientation not listed | 7 (2.3) | 2 (1.2) |
|  | Don't know | 11 (3.7) | 4 (2.4) |
|  | Prefer not to say | 5 (1.7) | 4 (2.4) |
| Employment Status, n (%) | Employed | 241 (80.6) | 144 (85.2) |
|  | Unemployed and actively seeking work | 7 (2.3) | 2 (1.2) |
|  | Not working and not actively seeking work | 39 (13.0) | 19 (11.2) |
|  | Prefer not to say | 12 (4.0) | 4 (2.4) |
| Medication Status, n (%) | Taking | 106 (35.5) | 65 (38.5) |
|  | Not taking | 193 (64.5) | 104 (61.5) |

The GAD-7 (screening only), PHQ-9 (screening only), WSAS, MPFI, SUS, SUTAQ and demographic data were collected via ieso’s secure care delivery platform used to routinely collect patient outcomes for NHS Talking Therapies. All clinical outcomes and demographic data for all control participants were also collected using this platform. GAD-7 and PHQ-9 check-ins throughout the program were collected using validated software within the smartphone app. Qualitative feedback and the UES were collected via Qualitrics^TM^. Safety endpoints were manually logged by research coordinators and clinicians following participant contact where events were reported (e.g. phone calls, clinical reviews, emails). All data were stored in a secure environment with restricted access, and extensive quality control was conducted to ensure data integrity.

### Description of Digital Intervention

The intervention consisted of a six-module digital program (‘ieso Digital Program’; software name: IDH-DP2-001) that used a conversational agent to guide participants through a pre-defined set of activities with human clinical oversight and user support. The program was intended as a first-line intervention for people primarily presenting with anxiety symptoms. The program was designed based on cognitive behavioral principles from traditional CBT and third wave approaches, such as ACT ^33,34^ (see Supplementary Table 1 for module details). All of the cognitive and behavioral processes, analogies and examples within the intervention were selected for their specificity in targeting symptoms of generalized anxiety.

The six modules consisted of an introduction module, three core modules, and two consolidation modules (Figure 2). The three core modules each consisted of three sessions that followed the pattern of i) learning, ii) activity, and iii) practice. The two consolidation modules consisted of two sessions. There were 16 sessions total. The introduction and consolidation modules consisted of sessions designed for onboarding and learning consolidation, respectively. All modules began with a symptom “check-in” consisting of the GAD-7 and PHQ-9 within the software immediately before the first session within that module. Sessions were made available on a timed schedule subject to completing the prior session (Figure 2).

**Figure 2.**
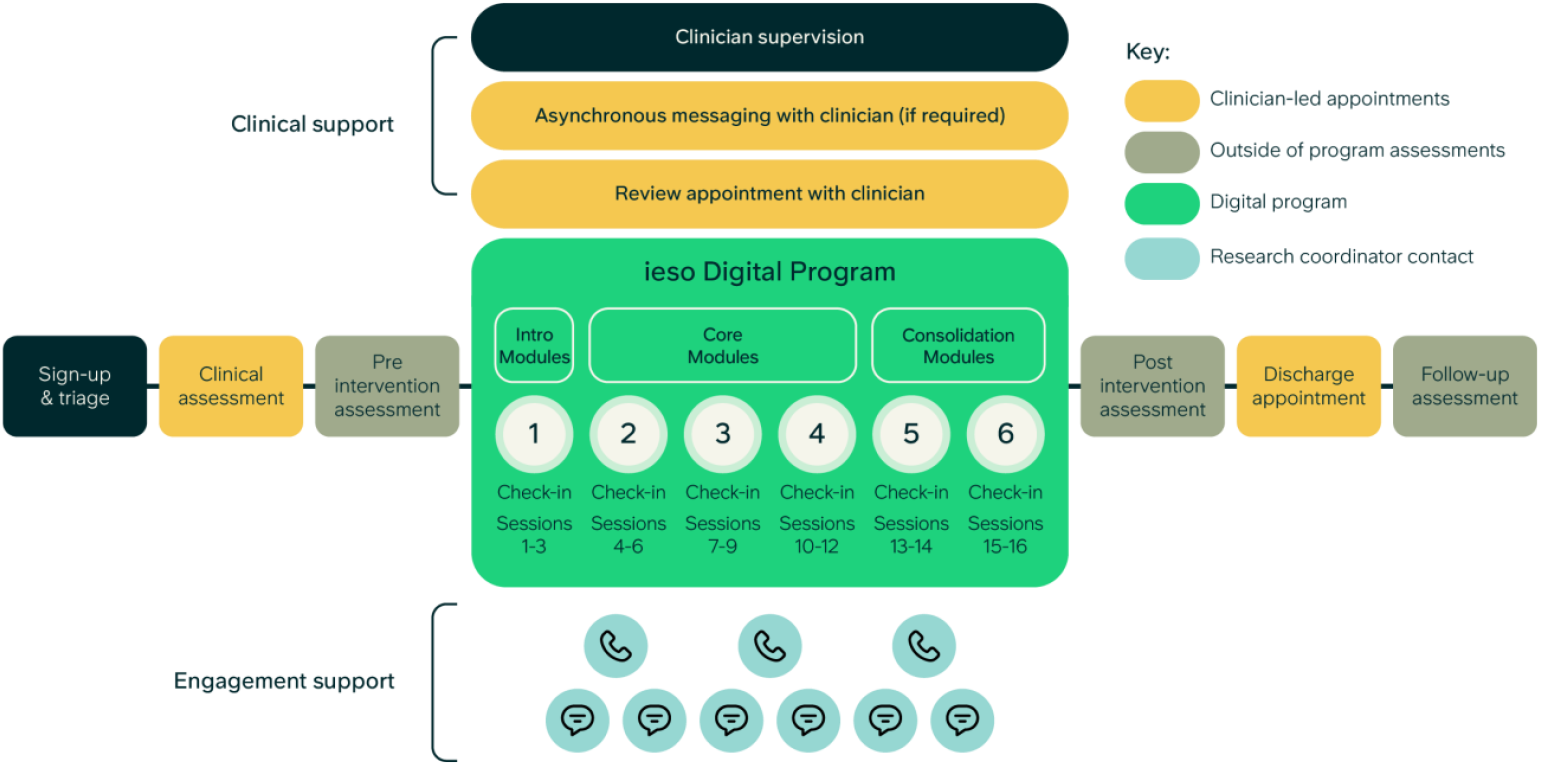
Schematic of ieso Digital Program with human clinical and user support service and study procedures. All participants received a clinical assessment prior to enrolment and were offered a discharge appointment with a clinician following the program. Clinicians were available via asynchronous messaging or for a review appointment whenever needed. All participants received email or SMS reminders and fortnightly check-in calls throughout the program to maximize engagement delivered via the research team. The ieso Digital Program included 6 modules with a total of 16 sessions. Each module started with a symptom check-in consisting of the GAD-7 and PHQ-9

Within each session, the software used a conversational agent to guide participants through a combination of videos, educational content, conversations, and worksheets written by accredited clinicians. The software used AI models for Natural Language Understanding, specific and tailored elements of Natural Language Generation and a dialogue management system. Part way through enrolment, with agreement from the overseeing NHS Research Ethics Committee, the software was updated to fix bugs, improve the user experience within the introductory module, and update select AI models. The final 60 participants enrolled were offered the updated software. Software version was controlled for in statistical analyses. The digital program was built in accordance with ISO 13485. Prior to the study, the program was registered as a UKCA marked Class 1 medical device.

### Human Support & Clinical Oversight

To ensure participant safety and maximize engagement and acceptability of the program, a dedicated human user and clinical support service was provided. Prior to enrolment, as part of the screening process, all participants received a standardized clinical assessment by a trained clinician with an accredited postgraduate qualification via typed modality. The clinician assessed the individual’s needs, determined if they were eligible for the study and obtained informed consent. Research coordinators provided fortnightly check-in calls to all participants throughout the program and sent weekly emails or SMSs to remind participants only if they deviated from the program schedule. Risk could be flagged through symptom monitoring of GAD-7 and PHQ-9 scores or through interaction with the research coordinators during check-in calls or ad hoc communication. Flagged risk was escalated to a clinician for review. Where appropriate the participant would then be contacted for further risk assessment by a clinician to ensure their safety. Participants could also request an appointment with a clinician at any point to discuss their journey, particularly if they were unsure the program was working for them. At the end of the study, all participants were offered a further discharge appointment with a study clinician to discuss the next steps for their care.

The support service and study procedures are illustrated in Figure 2. In total, delivering the intervention required an average of 97 minutes (1.6 hours) of clinician time (defined as time spent in sessions with participants) per participant. This included 299 assessments (mean 66 mins; range 31–105 mins), 47 review appointments (mean 32 mins; 14–60 mins across 46 participants) and 173 discharge appointments (mean 44 mins; range 13–76 mins).

### Participants

Adults with mild to severe symptoms of anxiety and a main presentation of Generalized Anxiety Disorder (GAD) were eligible as established through a standardized assessment conducted by a qualified clinician following the NHS TT manual^16^ and symptom severity on the GAD-7 scale. Individuals were invited to participate either following referral to ieso’s typed therapy service (either referred to ieso from the NHS Provider or via self-referral direct to ieso) or in response to online advertisements or email invitation through the NIHR BioResource for Translational Research (https://bioresource.nihr.ac.uk/). The program was not designed for individuals with a primary presenting problem of Depression, therefore participants with a PHQ-9 score ≥16 indicative of moderate to severe symptoms of Depression were signposted elsewhere for more appropriate support.

During the assessment, clinicians ensured all participants met the following eligibility criteria:

- over the age of 18 years at point of recruitment;
- GAD-7 total score > 7;
- PHQ-9 total score < 16;
- access to a smartphone and internet connection;
- registered with a General Practitioner in the UK;
- not currently receiving psychological therapy;
- suitable for CBT (excludes individuals with diagnosis of multiple disorders, psychotic or personality disorder, autism spectrum condition or intellectual disability)
- no diagnosis of an untreated mental health condition including substance misuse (except GAD or MDD)
- did not have PTSD, OCD or Panic Disorder;
- did not have a change in psychiatric medication in the past 1 month;
- did not display significant risk of harm to self, to others or from others (as established with the clinical assessment).

Any individuals who had previously participated in user research for the digital program were excluded. Participants were recruited between 10^th^ October 2023 and 2^nd^ February 2024. Financial incentive up to a total of *£*60 was provided in the form of vouchers based on study assessments and completion of modules within the digital program. For a sub-sample that participated in additional interviews, an additional *£*15 voucher per semi-structured interview was provided.

### Sample size

Previous studies have reported up to a 70% attrition rate when measuring engagement and adherence in mental health digital programs^35–37^, therefore we aimed to enroll 300 participants with the expectation of a 40-70% attrition rate, resulting in a final sample of 90-180 participants. A non-inferiority power analysis was conducted prior to retrospective analysis of external control data to estimate the total sample size needed to quantify clinical effectiveness (i.e. change in GAD-7 total score) compared to an active external control. Clinical effectiveness was defined as a change in GAD-7 score over either the course of six treatment sessions or until recovery was reached (if sooner than 6 sessions). A non-inferiority margin of a 1.8 change in GAD-7 total score was chosen based on previous literature^38–40^ (see Supplementary Methods for more details). Using data from 1489 patients being treated for GAD via typed-CBT, with at least six sessions or recovery, we estimated an expected standard deviation of GAD-7 change of 5.14. To estimate a sample size, we used the following equation: 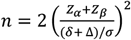 (see ^41^), where Zα and Z*β* are the standard normal scores for the one-sided significance level of 2.5% (1.96) and power of 90% (1.28) respectively, δ is the non-inferiority level 1.8 and σ is the standard deviation 5.14. A sample size of 172 was estimated for the study intervention to enable a non-inferiority analysis of clinical effectiveness compared to human-delivered care.

### Patient Public Involvement (PPI)

At ieso, experts-by-lived experience are involved in research and development work as members of a PPI panel and as partners advising on ongoing work. For this study, all participant facing documents were reviewed by members of the PPI panel. In addition, focus groups with members of the PPI panel during study conceptualization aimed to understand participant needs and expectations in the context of “keeping safe” whilst using the digital program, and helped developed recruitment marketing campaigns.

### External comparator data source for propensity-matched control groups

External comparator data were taken from two NHS TT service providers: i) ieso typed therapy data where a patient receives CBT through 1:1 communication with a qualified therapist using real-time text-based messaging; and ii) Dorset Healthcare University NHS Foundation Trust (DHC) delivering face-to-face routine therapy appointments. The information captured through the dataset of NHS TT is intended to support the monitoring of the implementation and effectiveness of national policy and legislation, policy development, performance analysis and benchmarking, national analysis and statistics, and national audit of NHS TT services. At registration, patients agree to the services’ terms and conditions, including the use of deidentified data for research and audit purposes, including academic publications or conference presentations. External control data were obtained from patients referred to: a) ieso’s typed therapy service between January 2022 and December 2023, and b) DHC between January 2017 and December 2021.

All control patients were propensity-matched to enrolled participants in the intervention group based on key predictors of treatment outcomes: baseline GAD-7 scores, baseline PHQ-9 scores, age, and the presence of a chronic physical health condition (yes/no/not known)^42^. Propensity-matching was conducted using the ‘*MatchIT*’ package^43^ in R with ‘nearest neighbor’ methodology (average treatment effect in treated patients), matching for propensity score on a one-to-one ratio. Comparator groups showed high similarity with the intervention sample (Supplementary Table 2). All propensity-matched control patients had a main presentation of GAD as established through the same standardized clinician assessment as the prospective participants (in line with the NHS TT manual). Treatment status and duration were matched as closely as possible and defined differently for the per-protocol and intention-to-treat samples outlined under *Statistical comparison to propensity-matched control groups*.

### Statistical methods

Analyses were conducted in R^44^. A statistical analysis plan was defined prior to final analyses being conducted.

#### Per-protocol vs. intention-to-treat samples

The per-protocol (PP) sample (n=169) was defined as participants who completed the minimum meaningful clinical dose of the program (MMCD) and the final post-intervention GAD-7 and PHQ-9 questionnaires. This dose was defined *a priori* by three accredited cognitive behavioral therapists who evaluated the content of the program to determine the amount of content required to deliver meaningful clinical improvement on the GAD-7 scale based on their clinical experience (mean experience of 14 years delivering psychological therapy). Based on this evaluation, the MMCD was defined as completing modules 1 to 3 in the digital program and the module 4 check-in.

The intention-to-treat (ITT) sample (n=299) included all participants who completed questionnaires at enrolment irrespective of adherence to the digital program except for one participant who requested that their data be deleted. Due to missing data for the pre-intervention WSAS (external recruits only), the ITT sample for all WSAS analyses was n=295.

#### Engagement and adherence analyses

Metrics of adherence were primarily assessed with descriptive statistics of in-software usage metrics: median and distribution of time spent in the digital program in hours, days since initialization of the program (defined based on the date that the software was downloaded); and proportion of participants completing each session, module, and check-in. An “engaged” patient is defined as an individual who has received the minimum amount of therapy such that pre- and post-treatment measures can be collected, and clinical outcomes estimated^16^. Here we used a comparable definition of engagement based on usage of the program (including time in the program, content delivered, and number of outcomes measured) defined as completing session 1 of module 2 in the program. This is in contrast to the MMCD definition which is defined based on both usage and expected improvement in symptoms.

#### Effectiveness analyses

Clinical effectiveness was quantified by calculating the change in anxiety symptoms, measured using the GAD-7, from baseline to final score, and estimating a within-subject effect size (Cohen’s *d*). The threshold for a clinically meaningful reduction in symptoms was defined as a change greater than the reliable change index of the GAD-7 scale (minimum of a 4-point reduction; Toussaint et al., 2020). A within-subjects effect-size for mean change in GAD-7 scores from post-intervention to one month follow-up was calculated to determine the short-term durability of any effects of the digital intervention. We also measured effectiveness by calculating the change in PHQ-9 and WSAS between baseline and final score, as well as between comparator groups. For the ITT sample, when calculating GAD-7 and PHQ-9 effectiveness, missing post-intervention scores were imputed using last observation carried forward, such that the final score collected prior to disengagement or withdrawal was used.

Clinical outcomes were calculated using the following definitions: a) improvement was defined as a reduction on the PHQ-9 or GAD-7 scales greater than or equal to the reliable change index ( ≥4 for GAD-7; ≥6 for PHQ-9) and no reliable increase on either measure; b) recovery was defined as reduction on both scales to below the clinical cutoff (GAD-7 score <8; PHQ-9 score <10); c) reliable recovery was defined as having both improved and recovered; d) responder rate was defined as an improvement of either ≥4 on the GAD-7 or ≥6 on the PHQ-9; and e) remission rate was defined as having either a final GAD-7 score <8 or final PHQ-9 score <10 for those only having started above the clinical cut-off. Definitions for improvement, recovery and reliable recovery are equivalent to those used in NHS TT^46^. Binary clinical outcomes were compared across groups using chi-squared tests. Bonferroni correction was used to account for multiple comparisons across related outcome metrics.

#### Regression models predicting adherence & effectiveness

To determine whether any demographic or study variables were associated with adherence or effectiveness, a series of regression analyses were conducted. All regression models included age, gender, highest qualification, employment status, religion, presence of a chronic physical health condition, ethnicity, reported disability, sexuality, baseline GAD-7 severity, software version, and enrolment path (referred to ieso’s typed therapy service or externally recruited) as predictors. Linear regression models were used to predict continuous dependent variables: i) number of sessions completed; ii) change in GAD-7 score from baseline to final score. A logistic regression model was used to predict non-adherence (i.e. participants who did not complete the necessary program sessions or study assessments to be in the PP sample; non-adherence coded as 1). Due to unequal sample sizes within demographic sub-categories (e.g. sexuality), groups were truncated to aid in the interpretability of findings and power of analyses.

Adherence was defined as the proportion of participants who completed each GAD-7 assessment (“session”) throughout their journey. For the ieso Digital Program group, each symptom check-in was at the beginning of each module within the program software (total 6 instances in program). For the therapy control groups, patients completed each GAD-7 assessment as part of each attended treatment session (either face-to-face or typed) up to 6 treatment sessions. Within NHS TT every attended treatment session includes a GAD-7 assessment. Sessions were aligned such that each symptom check-in within the digital program was associated with a treatment session for the control group. To determine if adherence across sessions differed between groups, a generalized linear model was used to test for a session-by-group interaction.

#### Statistical comparison to propensity-matched control groups

Three propensity-matched external control groups were created using real-world historic patient data (see *External comparator data source*) to compare the clinical effectiveness of the intervention to no intervention and standard of care. For the waitlist control only participants in the PP sample were matched (n=169) due to limited available data for matching. For the human-delivered therapy control groups all participants were matched (n=299).

The control groups consisted of:

i. *waiting controls (total available sample n=576)*; patients referred for typed-CBT with two GAD-7 scores between 4-10 weeks apart without having started treatment during that time (same sample used for PP and ITT analyses),
ii. *therapist delivered typed CBT (total available sample n=2*,*210)*; patients referred for typed-CBT with at least two scores on the GAD-7, who had completed a course of typed CBT - defined by the discharge code of ‘completed treatment’ - and discharged with a maximum of twelve treatment sessions (PP sample), or any patient who had entered treatment, regardless of completion (ITT sample), and
iii. *therapist delivered face-to-face CBT (total available sample n=753);* NHS TT patients referred to DHC who received face-to-face CBT and had a minimum of two and a maximum of twelve treatment sessions (PP sample), or any patient who attended treatment (ITT sample). Unlike the typed-CBT comparator, due to unavailability of discharge codes it was not possible to use the ‘completed treatment’ to define the PP sample for this group.

In line with the *a priori* defined statistical analysis plan, a superiority analysis was conducted to test the hypothesis that the clinical effectiveness of the intervention was greater than a propensity-matched waiting control group using a between-subjects t-test. A non-inferiority analysis was conducted to test the hypothesis that the clinical effectiveness of the intervention was not inferior to the effectiveness of typed CBT or face-to-face CBT in comparison to waiting-list. Within and between-subject effect sizes were also estimated for the change in total score on the PHQ-9 and the WSAS to estimate the effectiveness of the intervention on low mood and work and social functioning relative to the waiting control

## Results

The final sample for analysis included 299 participants of whom 80% were female (n=240) with a mean age at baseline of 39.8 years (range: 18 – 75 years). Table 1 provides an overview of demographics and baseline severity for participants in the intervention group for both the ITT and PP samples.

### Engagement and adherence

Participants (n=299) completed a median of 6.1 hours of program interaction over 53.1 days. This was higher for the PP sample in which participants completed a median of 8.7 hours over 59.6 days. In total, 232 participants (78%) were engaged in the program (i.e. completed session 1 of module 2) involving a median of 2 hours interacting with the program content over 14 days. Out of those engaged participants, 78% (n=180) reached the minimum meaningful clinical dose (i.e. completing up to check-in 4 out of 6 in the program). The overall study attrition rate (defined as the proportion of participants who did not complete the final study questionnaires) was 32%. Descriptive statistics of engagement with the program across modules are outlined in Table 2.

**Table 2.** Engagement metrics for the digital program.

|  | N | Median time since initialization (days) | Median time interacting in program (hours) |
| --- | --- | --- | --- |
| Per-protocol sample total | 169 | 59.6 | 8.7 |
| Intention-to-treat sample total | 299 | 53.1 | 6.1 |
| Engaged sample total (up to module 2 session1) | 232 | 14.0 | 2.0 |
| *All participants by milestone* |  |  |  |
| Module 1 check-in | 284 | 0.0 | 0.03 |
| Module 2 check-in | 240 | 13.6 | 1.5 |
| Module 3 check-in | 209 | 23.9 | 2.7 |
| Module 4 check-in | 180 | 35.0 | 3.8 |
| Module 5 check-in | 138 | 42.9 | 5.0 |
| Module 6 check-in | 113 | 49.5 | 5.4 |

Median days since initialization was calculated as number of days since software download at onboarding for each sample: PP, ITT and the engaged sample (i.e. those who completed up to session 1 of module 2). Metrics are also shown for each symptom check-in at the beginning of each module.

To determine if adherence across sessions differed between groups, adherence rates were compared using a session-by-group interaction. There was a significant effect of session number (*t* = –6.4, *p* < .001), but no significant session-by-group interaction for face-to-face therapy (*p* = 0.18) or typed-therapy (*p* = 0.76) indicating no difference in adherence rates across groups (Figure 3; model output in Supplementary Table 3).

**Figure 3.**
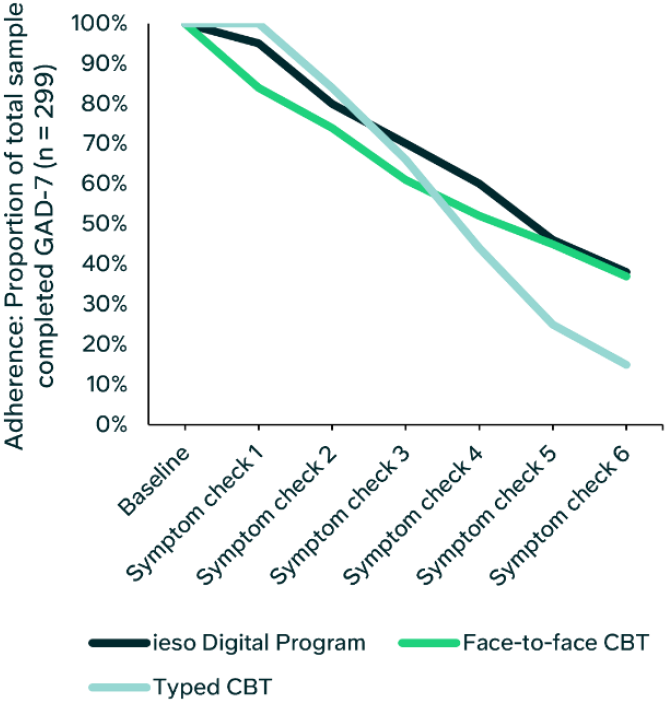
Adherence with program progression overlaid with adherence across therapy sessions for the control groups. For each group, adherence was defined based on the proportion of participants who completed each GAD-7 assessment (“symptom check”) throughout their journey. Baseline was 100%, i.e. all participants/patients attended a clinical assessment and had a baseline GAD-7 score. For the ieso Digital Program group, each symptom check-in was at the beginning of each module within the program software (total 6 instances in program). To complete each symptom check-in within the program, participants had to finish the previous module. For the therapy control groups, patients completed each GAD-7 assessment as part of each attended treatment session (either face-to-face or typed) up to 6 treatment sessions. Within NHS TT every attended treatment session includes a GAD-7 assessment. Adherence rates across sessions were not significantly different between groups(Supplementary Table 3).

To investigate potential drivers of program adherence, demographic and study factors were associated number of completed sessions in the program. Only age was significantly associated with adherence such that older participants were more likely to complete more sessions in the program (linear regression: F(25, 273) = 1.3, *p* = .13, adjusted R^2^ = 0.03; age effect: *b* = 0.11, SE = 0.04, *t* = 2.65, *p* = .009; Supplementary Table 4). Older participants were also more likely to be included in the PP sample (Supplementary Table 5).

### Clinical Effectiveness

#### Anxiety symptoms

On average, across the intervention sample, there was a large, clinically meaningful reduction in anxiety symptoms from baseline to final score (PP: mean GAD-7 change = –7.4, 95% CI [–8.1, –6.7], *d* = 1.6; ITT: mean GAD-7 change = –5.4 [–6.0, –4.8], *d* = 1.1; Figure 4, Table 3). This reduction was significantly greater than the waiting control (mean GAD-7 change = –1.9 [–2.5, –1.3]; PP between-subject effect: *p* <.001, *d* = 1.3; ITT between-subject effect: *p* <.001, *d* = 0.8), and statistically non-inferior to the face-to-face therapy control (PP: mean GAD-7 change = –6.4 [–7.0, –5.8], non-inferiority effect *p* <.001; ITT: mean GAD-7 change = –6.0, [–6.6, –5.5], non-inferiority effect *p* = .002). For the typed-therapy control, the intervention was significantly non-inferior for the PP sample (mean GAD-7 change: –7.5, [–8.0, –7.0]; non-inferiority *p* <.001), and for the ITT sample the effect was approaching significance (mean GAD-7 change = –6.6 [– 7.1, –6.1], *p* = .06). Clinical outcomes were consistently greater for the digital program compared to the waiting control and similar across the active control arms for the PP sample. Outcomes are reported in Supplementary Table 6 & 7.

**Table 3.**
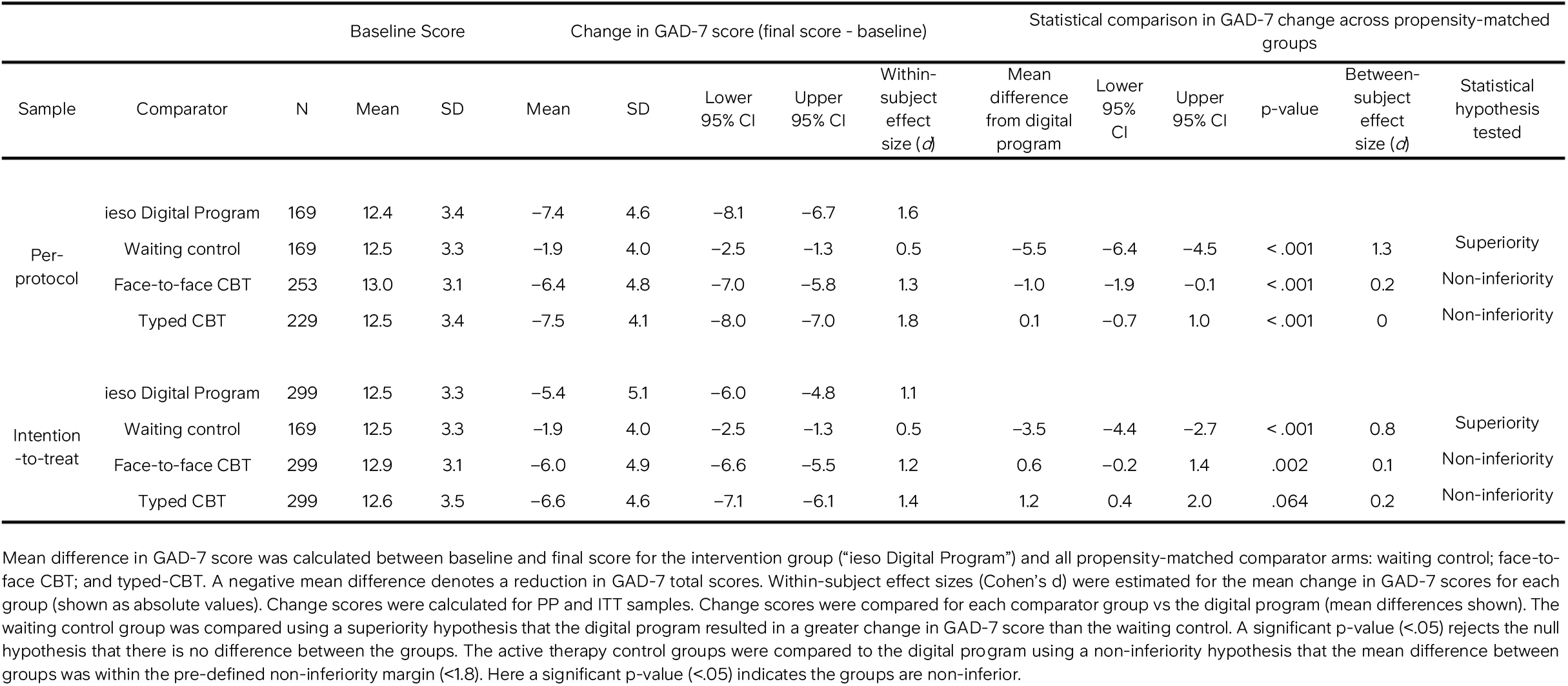
Change in GAD-7 score from baseline to final score for the intervention sample and propensity-matched control groups.

**Figure 4.**
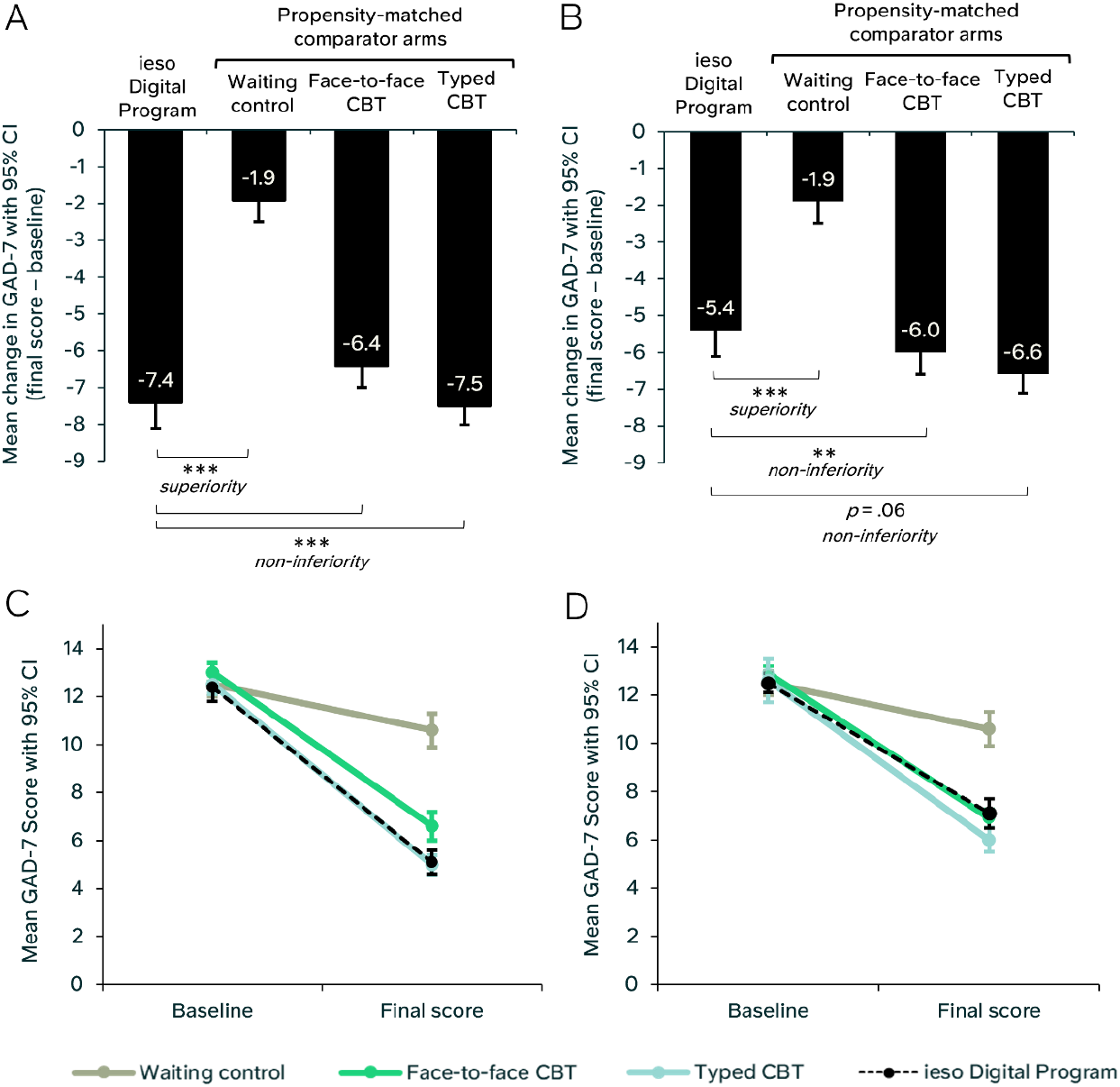
Change in anxiety symptoms from baseline to final score for the intervention sample and propensity-matched control groups. A) Mean change (final score – baseline) in GAD-7 scores for the PP sample (n=169), propensity-matched waiting control group, face-to-face CBT group, and typed CBT group. B) Mean change in GAD-7 scores for the ITT sample (n=299) and all control groups. C) Mean GAD-7 scores at baseline and final score with 95% confidence intervals for the PP sample (n=169) and all control groups. D) Mean GAD-7 scores at baseline and final score with 95% confidence intervals for the ITT sample (n=299) and all control groups. ^***^ = *p* < .001, ^**^ = *p* <.005

The trajectory for mean reduction in anxiety symptoms was steeper following the earlier program modules (Figure 5; Supplementary Table 8). When stratified by baseline GAD-7 severity into mild, moderate, and severe groups, the severe group showed the greatest reduction in anxiety symptoms (PP (n=48): mean change on GAD-7 = –10.7 [–12.3, –9.2], *d* = 2.0; ITT (n=87): mean change on GAD-7 = –7.9, [–9.2, –6.6], *d* = 1.3; Supplementary Table 9).

**Figure 5.**
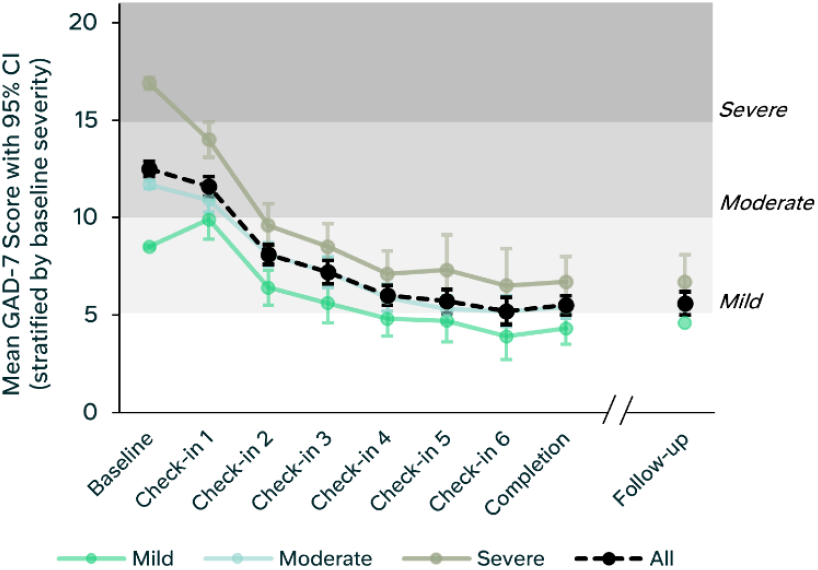
Mean reduction in anxiety symptoms across digital program. Mean GAD-7 score for each time-point for all participants that completed the questionnaires at each time-point. Trajectories split by GAD-7 baseline severity: mild, moderate and severe (Supplementary Table 8).

By the end of the program, the moderate and severe groups showed a mean GAD-7 score in the mild range. The clinical effect was sustained at one-month follow-up (Figure 5). Between final score and one month follow-up, there was no change in GAD-7 mean score for the PP sample (n=166; mean change = 0.0 [–0.4, 0.5]) and ITT sample (n=210; mean change = 0.0 [–0.5, 0.4]; Supplementary Table 8).

The associations between participant demographics, study factors and change in GAD-7 score were explored with a linear regression: F(25, 273) = 3.31, p< .001, adjusted R^2^ of 0.16. Greater reductions in GAD-7 scores were associated with higher baseline GAD-7 scores (*b* = 0.69, SE = 0.09, *t* = 7.46, p< .001), and higher baseline age (*b* = 0.08, SE = 0.03, *t* = 3.0, *p* = .003) (Supplementary Table 10), such that more severe, and older participants saw a larger change in GAD-7 score.

#### Mood symptoms

As intended, given the specificity of the program for targeting symptoms of generalized anxiety, there was a statistically significant yet smaller effect for low mood symptoms as measured with the PHQ-9 (PP: mean PHQ-9 change = –3.1 [–3.8, –2.4], *d* =0.7; ITT: mean PHQ-9 change = –1.6 [–2.1, –1.1], *d* = 0.3) (Table 4). This mean change was significantly greater than the mean change in the waiting control group for the PP sample (mean PHQ-9 change = –1.0, [–1.5, –0.4], between-subject effect, *p* < .001, *d* = 0.5), but not for the ITT sample (*p* = .11 *d* = 0.1). Despite this, PHQ-9 remission rate (based on n=116 above the clinical cut-off at baseline) was 67% for the ITT sample (Supplementary Table 7). Participants with severe and moderate baseline GAD-7 scores experienced the largest improvement in PHQ-9 scores (Supplementary Table 9). There was minimal mean change in scores between post intervention and follow-up for both PP and ITT samples (PP mean difference = 0.5 [0.0, 1.0]; ITT mean difference = 0.4 [–0.1, 0.9]) (Supplementary Table 11).

**Table 4.**
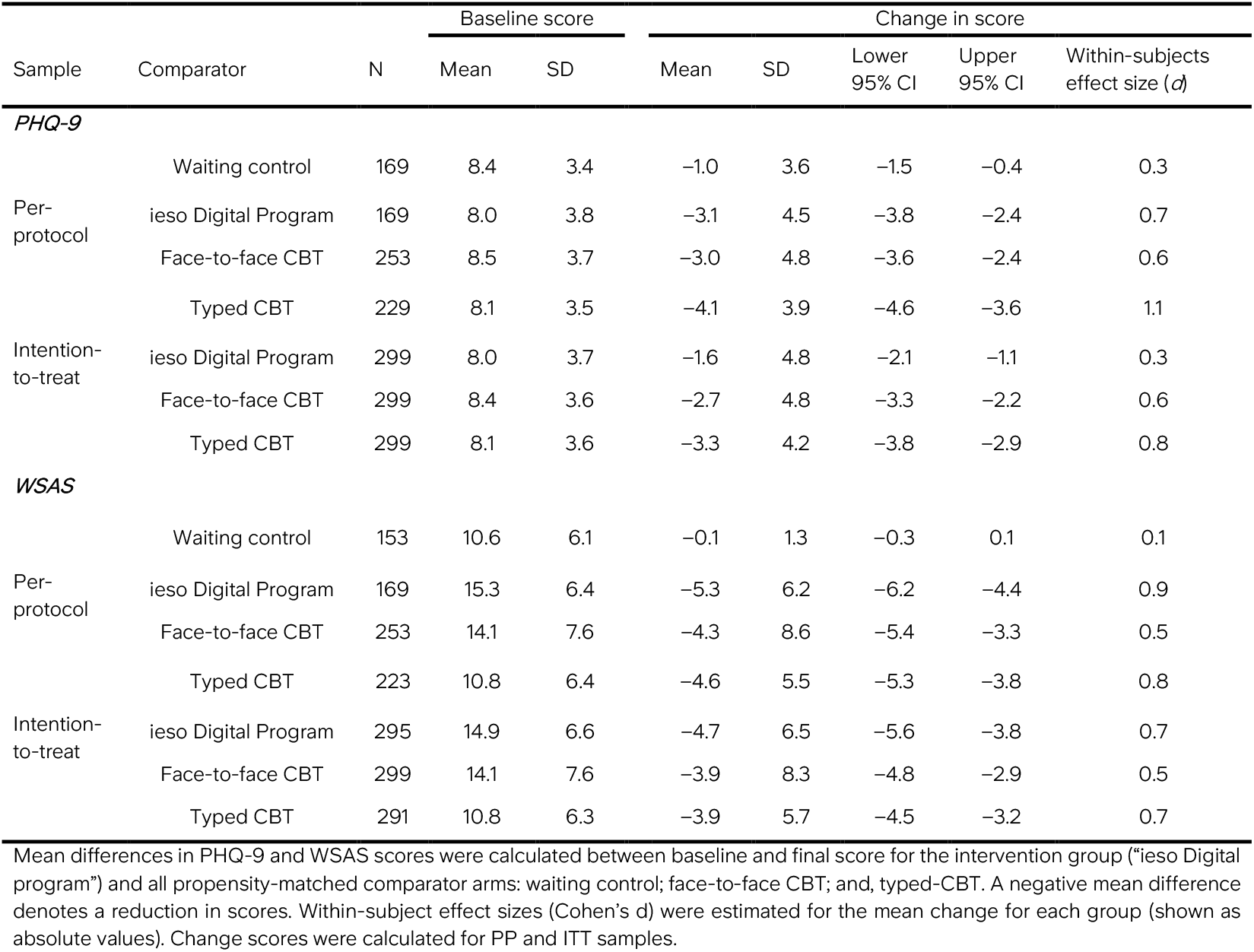
Change in PHQ-9 and WSAS score from baseline to final score for all groups.

#### Work and social functioning

There was a significant improvement in work and social functioning measured using the WSAS from baseline to final score for the intervention group (PP: mean WSAS change = –5.3 [–6.2, –4.4], *d* = 0.9 ; ITT (n=295): mean WSAS change = –4.7 [–5.6, –3.8], *d* = 0.7) (Table 4). This mean change was significantly greater than the mean change in the waiting control group (mean WSAS change = –0.1; PP between-subject effect *p* < .001, *d* = 1.2; ITT between-subject effect, *p* < .001, *d* = 0.8). The largest changes in functioning were for severe and moderate anxiety groups (Supplementary Table 9).

### Safety

The digital program was well tolerated, and no serious adverse events were identified during the study. There was one report of migraine and two reports of insomnia. There were 10 software deficiencies that occurred (for 7 participants; 90% prior to the software update) for reasons such as technical issues or difficulties with the conversational agent understanding users. In all instances participants were offered an appointment to discuss any potential impact of this on their mental health and reminded of their right to withdraw. These instances resulted in one active participant withdrawal. Across the study, 10 participants were withdrawn by a study clinician following a conversation between the participant and a study clinician. These withdrawals were linked to the study exclusion criteria and suitability for the program rather than the safety of the intervention.

## Discussion

This study demonstrates that an evidence-based, human-supported digital intervention for adults with mild, moderate and severe anxiety produced a large clinically meaningful reduction in anxiety symptoms significantly greater than a propensity-matched waiting control and non-inferior to real-world human delivered therapy. Engagement with the program was high and participants adhered to the intervention at a similar rate to the external therapy control groups. The intervention achieved comparable reduction in anxiety symptoms to human-delivered care with significantly reduced clinician time. By integrating technology and human support, this study demonstrates the potential to expand global access to high-quality, effective mental healthcare.

The large clinical effect of the digital intervention across participants with moderate or severe symptoms highlights the clinical value of the combined program content and human support. Here, the PP (*d* = 1.3) and ITT (*d* = 0.8) effect sizes relative to waitlist are larger than the pooled effect size reported in a recent meta-analysis of digital interventions without any blended-care component (n comparisons = 96, *g* = 0.26)^14^. Unlike the PP sample which is designed to demonstrate the clinical effectiveness of an intervention when the intervention is adhered to, the ITT sample provides an estimate of effectiveness more reflective of the real-world context by accounting for disengagement. The large ITT effect was significantly non-inferior to face-to-face therapy, and approaching significance for non-inferiority to typed-therapy (*p* = 0.06). Human-delivered care enables greater flexibility to respond to patient concerns and adapt content compared to a digital program. However, the comparable clinical effects and adherence rates across groups, particularly for the PP sample, indicates the potential of this digital intervention to significantly impact real-world patient outcomes.

It is important to note that high relapse and recurrence rates have implications for both patient quality of life and economic healthcare costs, therefore ensuring effects are durable is imperative^47–49^. Incorporating cognitive and behavioral principles into daily life through practical exercises can enable meaningful behavioral change that persists beyond treatment end. Here, both the persistent clinical effect at one month follow-up, and the significant improvement in the impact of anxiety on participants’ day-to-day functioning (as measured with the WSAS) highlights the potential of the intervention to instigate long-lasting behavioral change. Retrospective analysis of recurrence data from electronic health records is needed to accurately measure the persistence of the clinical effect in the real world over a longer follow-up time-period.

The engagement rate of the digital program (78%) and time to reach “engaged” (∼2 hours of program interaction over 2 weeks) is comparable to engagement rates and time in therapy observed in NHS TT services for treatment of GAD (70%; 2022-2023)^50^. Adherence rates across groups in the study were also similar. Average program interaction time (median 6.1 hours) across the ITT sample was greater than that reported for similar app-based interventions (e.g. median 3.4 hours)^51^, indicating high engagement with the program. Study attrition (32%) was higher than previous reports from studies of conversational agent-delivered mental health interventions (22%)^52^, yet similar to real-world global treatment drop-out rates (∼20-40%)^53,54^. This may be due to the pragmatic design of the study: 30% of the sample recruited through ieso’s therapy referrals could choose to withdraw at any time and immediately access 1:1 human-delivered therapy; and participants had the option to discuss their progress or any issues with the clinical team at any point. These factors could have increased withdrawal rates more than previous studies, but more readily reflect real-world patient choice and clinical decision-making.

To our knowledge, this study is the first to compare the effectiveness of a digital intervention to standard of care using external propensity-matched comparator groups from real-world patient data. There is increasing acceptability for the use of externally controlled clinical trials^55–58^ made possible by the availability of large-scale, standardized datasets. Generating external comparator groups reduces patient burden, study costs, and avoids delaying treatment for the comparator group receiving no intervention^59^. Here control groups were of high quality according to Thorlund and colleagues validity criteria^60^: 1) control data were from real-world NHS TT services with the same clinical assessment, outcomes and data collection procedures (in accordance with the NHS TT manual) as the prospective participants; 2) controls selected had the highest similarity in baseline characteristics to the intervention group due to the propensity-matching procedure (Supplementary Table 2); and 3) an a priori power analysis ensured samples were of adequate size to test for non-inferiority. However, creating standard of care control arms that are directly comparable to a novel intervention is difficult due to differences in how to define comparable doses, treatment completion and account for study-specific assessments. Moreover, the lack of randomization in the current study means selection bias and unmeasured variable effects are not controlled for. Randomization is the gold-standard for measuring efficacy in clinical trials as it ensures results are not biased and increases confidence that the outcomes measured are attributable to the intervention itself. However, effect sizes often do not generalize to the real-world where outcomes *are* biased by a patient’s preference over their treatment. This pragmatic study design may therefore more readily reflect effectiveness in a real-world context and offers a method for estimating effectiveness more quickly and cost-effectively reducing time from intervention development to patient benefit.

The clinical effect and engagement rate reported in the current study could have been driven by a combination of the three key features of the digital intervention: i) a curated and structured evidence-based program, ii) a conversational agent to deliver the program content, and iii) a human user and clinical support model akin to standard healthcare delivery. First, the structured evidence-based program was curated by a team of accredited cognitive behavioral therapists with an average of 14 years direct clinical experience. The program used principles from traditional CBT^22^ including third wave approaches, such as ACT. This approach encourages individuals to accept their thoughts and feelings while committing to actions aligned with their values. There is a growing body of evidence indicating that ACT demonstrates comparable effectiveness to other forms of CBT for anxiety disorders^61–63^, and has been shown to be acceptable and engaging within a digital program for GAD^64,65^.

Second, a conversational agent was used to personalize the content delivery and enhance engagement. Despite rapid growth in the development of AI conversational agents, use of this technology remains rare in digital mental health interventions, with only ∼5% using this technology^14^. The majority of these systems employ a tree-based dialogue approach, where natural language processing analyzes user input, and responses are selected from a predefined set of pre-written answers. However, previous research has shown users find this frustrating, particularly when it feels the agent does not understand them^66,67^. Recent advances in the development of large language models now make it possible to flexibly generate personalized language for a more engaging user experience. In the current study, the digital program primarily used a tree-based dialogue system with controlled use of natural language generation in specific instances to enhance engagement. Increased use of generative technology and reduced reliance on tree-based approaches will continually improve the capability of conversational agents to create a personalized and engaging experience. However, allowing fully autonomous language generation within the context of mental health, where patient problems can be nuanced, complex and require the consideration of social and cultural contexts, poses a high risk for patient harm and misuse^68^. Stringent validation of these new AI technologies with a phased roll out alongside human oversight will be essential to ensure patient safety^69^.

Finally, a ‘blended’ design of human support and conversational technology has been suggested to be key for maximizing real-world engagement^52^. Previous research has highlighted lack of trust, lack of user-centric design, privacy concerns, poor usability, and being unhelpful in emergencies as key drivers of poor engagement with digital interventions^12^. To address these concerns, we mirrored a real-world treatment model including user support services, clinician referral to the program, proactive symptom monitoring and clinician availability for collaborative decision-making with each participant. This service created a credible and trustworthy patient experience that we believe positively impacted patient outcomes. Although this study was not designed to demonstrate the economic value of the intervention, the average clinician time spent per participant was <2 hours, which is significantly lower than current standards of care globally: approximately 4 times less than an average episode of treatment in the UK for GAD (∼8 appointments between 45-60 mins; NHS Digital 2021-2022)^50^ and ∼approximately 8 times less globally (∼15 appointments; mean across reported naturalistic studies in ^70^). This new model, combining an AI-driven program with clinical support, allows the current, limited supply of trained therapists to help more people than current standards of care.

Limitations of the current study include the use of compensation for time for those who volunteered to participate, and the selection of a sample with limited low mood symptoms. In particular, in line with the study exclusion criteria, individuals with severe depression symptoms were not included. Nevertheless, the propensity-matching across groups controls for this, i.e. all groups included patients with similar baseline anxiety and depression symptoms. Differences in PP sample sizes across the control groups were likely driven by the definition of PP in each context rather than engagement, given similar adherence rates across the groups. Defining a comparable PP sample across groups is challenging due to differences in dose intensity, delivery mechanism and data collected, as well as significant variation across patients in both clinical presentation of generalized anxiety and response to treatment. The PP samples for the therapy control groups were based on completed episodes of care, therefore were agnostic of therapy dose and would have included those who received a low number of sessions and recovered quickly. Those individuals would not have been included in the intervention PP sample which was conservatively defined based on minimum program interaction over 9 weeks. This was study was not randomized and relied on patient-reported outcomes, therefore a prospective randomized clinical trial with clinician reported outcomes will be useful to confirm clinical efficacy of the intervention.

There were also limitations in terms of the diversity of the intervention sample, with enrolled participants predominantly white, highly educated, and female. This sample is reflective of the typical profile of GAD patients in the UK and US^50,71^. Although we attempted to increase diversity in this sample through focused marketing campaigns, these efforts were not successful. Needs differ across individuals, conditions and contexts, and a greater understanding of the barriers to research participation is required to fully understand these needs, particularly where groups have been systematically excluded from research, and where there is stigma around mental health. Increasing access to mental health support could play a substantial role in addressing unmet need in underserved groups, therefore future work will aim to evidence the inclusivity of this digital intervention and its potential to counter existing health inequalities.

In conclusion, this study demonstrates that a digital intervention, designed for adults with symptoms of generalized anxiety, produces comparable outcomes to human-delivered CBT while significantly reducing the required clinician time. This result indicates the potential for digital interventions to provide high quality, evidence-based care at scale to address unmet need worldwide. As AI technologies rapidly progress, it is evident that generative dialogue systems that emulate creative and flexible human language will soon be widely accessible. This accessibility will radically change how individuals seek mental health support. Our responsibility lies in leveraging these advances, addressing the ethical and social challenges inherent with AI, and combining the best of technology with the best of clinical care to increase access to effective, safe and engaging mental health support for everyone. Rigorous evidence, particularly to understand the optimal blend of human and computer support for different individuals, will be key to accelerate precision treatment, maintain scalability, maximize uptake and adherence, and integrate digital interventions into health systems.

## Acknowledgements

We extend our gratitude to the patients who participated in the study and to the dedicated clinicians and support staff involved. We would like to thank Gerald Chan, Stephen Bruso, Andy Richards, Ann Hayes, David Icke, Michael Black, Clare Hurley, Florian Erber, Richard Marsh, Sam Williams, Jo Parfrey for their support and encouragement. We are grateful to Prof Thalia Eley for introducing us to NIHR BioResource. We thank NIHR BioResource volunteers for their participation, and gratefully acknowledge NIHR BioResource centers, NHS Trusts and staff for their contribution. We thank the National Institute for Health and Care Research, NHS Blood and Transplant, and Health Data Research UK as part of the Digital Innovation Hub Program. The views expressed are those of the author(s) and not necessarily those of the NHS, the NIHR or the Department of Health and Social Care. We thank Dorset Healthcare University NHS Foundation Trust (DHC) for providing external data for comparison.

## Funding

This research was funded by ieso Digital Health Ltd.

## Competing Interests

Chief Investigator (EMa) and other investigators (CEP, EMi, GW, MPE, EC, SL, AS, CH, JY, MB, LM, SM, RC, VT, AC, AW, AB) are employees of ieso Digital Health Limited (the company funding this research) or its subsidiaries. None of these authors had a direct financial incentive related to the results of this study or the publication of the manuscript.

## Author contributions

CEP, EMa, AW & AB conceptualized the study. CEP and EMi drafted the paper. GW, EMi, MPE, EC, MB, AS & AC contributed to data analyses and interpretation. SL, JY, CH, EMa & CEP conducted the study. All authors contributed to the interpretation of results and paper revision, and approved the final version.

## Data availability

Owing to the potential risk of patient identification, and following data privacy policies at ieso and DHC, individual-level data are not available. Aggregated data are available upon request, subject to a data-sharing agreement with ieso and DHC. Data requests should be sent to the corresponding author and will be responded to within 30 days.

## Notes

### Clinical Trial

ISRCTN ID: 52546704

### Author Declarations

NHS Research Ethics Committee (REC) West of Scotland REC 4 gave ethical approval for this research (IRAS ID: 327897)

### Summary of Updates

Table 3 updated to include between-subject effects; Discussion revised to increase clarity around limitations; Supplementary table 3 updated to include more details around statistical findings; Supplementary table 6 expanded into two tables and post-hoc chi-squared tests added

## References

1. World Health Organisation. Mental Health | Key facts. 2022. Accessed June 24, 2024. https://www.who.int/news-room/fact-sheets/detail/mental-disorders#:~:text=In%202019%2C%201%20in%20every%208%20people%2C%20or,anxiety%20and%20depressive%20disorders%20the%20most%20common%20%281%29.

2. Alonso J, Liu Z, Evans-Lacko S, et al. Treatment gap for anxiety disorders is global: Results of the World Mental Health Surveys in 21 countries. Depress Anxiety. 2018;35(3):195–208. doi:10.1002/da.22711

3. Our World in Data. Psychiatrists per 100,000 people. 1. March 21, 2024. Accessed July 3, 2024. https://ourworldindata.org/grapher/psychiatrists-working-in-the-mental-health-sector

4. Health Resources & Services Administration. Health Workforce Shortage Areas. February 7, 2024. Accessed July 3, 2024. https://data.hrsa.gov/topics/health-workforce/shortage-areas

5. Roland J, Lawrance E, Insel T, Christensen H. THE DIGITAL MENTAL HEALTH REVOLUTION TRANSFORMING CARE THROUGH INNOVATION AND SCALE-UP: WISH 2020 Forum on Mental Health and Digital Technologies.; 2020.

6. Clay R. Mental health apps are gaining traction. January 1, 2021. Accessed July 3, 2024. https://www.apa.org/monitor/2021/01/trends-mental-health-apps

7. Torous J, Roberts LW. Needed innovation in digital health and smartphone applications for mental health transparency and trust. JAMA Psychiatry. 2017;74(5):437–438. doi:10.1001/jamapsychiatry.2017.0262

8. Lattie EG, Stiles-Shields C, Graham AK. An overview of and recommendations for more accessible digital mental health services. Nature Reviews Psychology. 2022;1(2):87–100. doi:10.1038/s44159-021-00003-1

9. Borghouts J, Eikey E, Mark G, et al. Barriers to and facilitators of user engagement with digital mental health interventions: Systematic review. J Med Internet Res. 2021;23(3). doi:10.2196/24387

10. M. Ng M, Firth J, Minen M, Torous J. User engagement in mental health apps: A review of measurement, reporting, and validity. Psychiatric Services. 2019;70(7):538–544. doi:10.1176/appi.ps.201800519

11. Michie S, Yardley L, West R, Patrick K, Greaves F. Developing and evaluating digital interventions to promote behavior change in health and health care: Recommendations resulting from an international workshop. J Med Internet Res. 2017;19(6). doi:10.2196/jmir.7126

12. Torous J, Nicholas J, Larsen ME, Firth J, Christensen H. Clinical review of user engagement with mental health smartphone apps: Evidence, theory and improvements. Evid Based Ment Health. 2018;21(3):116–119. doi:10.1136/eb-2018-102891

13. Tafradzhiyski N. Mobile App Retention. Business of Apps.

14. Linardon J, Torous J, Firth J, Cuijpers P, Messer M, Fuller-Tyszkiewicz M. Current evidence on the efficacy of mental health smartphone apps for symptoms of depression and anxiety. A meta-analysis of 176 randomized controlled trials. World Psychiatry. 2024;23(1):139–149. doi:10.1002/wps.21183

15. David D, Cristea I, Hofmann SG. Why cognitive behavioral therapy is the current gold standard of psychotherapy. Front Psychiatry. 2018;9(JAN). doi:10.3389/fpsyt.2018.00004

16. The National Collaborating Centre for Mental Health. The Improving Access to Psychological Therapies Manual The Improving Access to Psychological Therapies Manual - Appendices and Helpful Resources.; 2018.

17. Ewbank MP, Cummins R, Tablan V, Catarino A, Buchholz S, Blackwell AD. Understanding the relationship between patient language and outcomes in internet-enabled cognitive behavioural therapy: A deep learning approach to automatic coding of session transcripts. Psychotherapy Research. Published online 2020:1–13. doi:10.1080/10503307.2020.1788740

18. Ewbank MP, Cummins R, Tablan V, et al. Quantifying the Association between Psychotherapy Content and Clinical Outcomes Using Deep Learning. JAMA Psychiatry. 2019;77(1):35–43. doi:10.1001/jamapsychiatry.2019.2664

19. Huckvale K, Venkatesh S, Christensen H. Toward clinical digital phenotyping: a timely opportunity to consider purpose, quality, and safety. NPJ Digit Med. 2019;2(1). doi:10.1038/s41746-019-0166-1

20. Catarino A, Harper S, Malcolm R, et al. Economic evaluation of 27,540 patients with mood and anxiety disorders and the importance of waiting time and clinical effectiveness in mental healthcare. Nature Mental Health. 2023;1(9):667–678. doi:10.1038/s44220-023-00106-z

21. Taylor HL, Menachemi N, Gilbert A, Chaudhary J, Blackburn J. Economic Burden Associated with Untreated Mental Illness in Indiana. JAMA Health Forum. 2023;4(10):E233535. doi:10.1001/jamahealthforum.2023.3535

22. Fenn K, Byrne M. The key principles of cognitive behavioural therapy. InnovAiT: Education and inspiration for general practice. 2013;6(9):579–585. doi:10.1177/1755738012471029

23. Wilson K, Hayes S, Strosahl K. Acceptance and Commitment Therapy: An Experiential Approach to Behavior Change. Guilford Press; 2003.

24. Gilbody S, Brabyn S, Lovell K, et al. Telephone-supported computerised cognitive-behavioural therapy: REEACT-2 large-scale pragmatic randomised controlled trial. British Journal of Psychiatry. 2017;210(5):362–367. doi:10.1192/bjp.bp.116.192435

25. Schulz KF, Altman DG, Moher D. CONSORT 2010 Statement: Updated guidelines for reporting parallel group randomised trials. BMJ (Online). 2010;340(7748):698–702. doi:10.1136/bmj.c332

26. Spitzer RL, Kroenke K, Williams JBW, Löwe B. A Brief Measure for Assessing Generalized Anxiety Disorder The GAD-7. Arch Intern Med. 2006;166:1092–1097.

27. Kroenke K, Spitzer RL. The PHQ-9: A New Depression Diagnostic and Severity Measure. Psychiatr Ann. 2002;32(9):509–515. doi:10.3928/0048-5713-20020901-06

28. Mundt JC, Marks IM, Shear MK, Greist JH. The Work and Social Adjustment Scale: A simple measure of impairment in functioning. British Journal of Psychiatry. 2002;180(MAY):461–464. doi:10.1192/bjp.180.5.461

29. Rolffs JL, Rogge RD, Wilson KG. Disentangling Components of Flexibility via the Hexaflex Model: Development and Validation of the Multidimensional Psychological Flexibility Inventory (MPFI). Assessment. 2018;25(4):458–482. doi:10.1177/1073191116645905

30. O’Brien HL, Cairns P, Hall M. A practical approach to measuring user engagement with the refined user engagement scale (UES) and new UES short form. International Journal of Human Computer Studies. 2018;112:28–39. doi:10.1016/j.ijhcs.2018.01.004

31. Brooke J. Usability Evaluation in Industry. 1st Edition.; 1996. doi:10.1201/9781498710411-35

32. Hirani SP, Rixon L, Beynon M, et al. Quantifying beliefs regarding telehealth: Development of the Whole Systems Demonstrator Service User Technology Acceptability Questionnaire. J Telemed Telecare. 2017;23(4):460–469. doi:10.1177/1357633X16649531

33. Hayes S, Follette V, Linehan M. Mindfulness and Acceptance: Expanding the Cognitive-Behavioral Tradition. Guilford Press; 2011.

34. Berg H, Akeman E, McDermott TJ, et al. A randomized clinical trial of behavioral activation and exposure-based therapy for adults with generalized anxiety disorder. Journal of Mood and Anxiety Disorders. 2023;1:100004. doi:10.1016/j.xjmad.2023.100004

35. Beatty C, Malik T, Meheli S, Sinha C. Evaluating the Therapeutic Alliance With a Free-Text CBT Conversational Agent (Wysa): A Mixed-Methods Study. Front Digit Health. 2022;4. doi:10.3389/fdgth.2022.847991

36. Boucher E, Honomichl R, Ward H, Powell T, Stoeckl SE, Parks A. The Effects of a Digital Well-being Intervention on Older Adults: Retrospective Analysis of Real-world User Data. JMIR Aging. 2022;5(3). doi:10.2196/39851

37. Cliffe B, Croker A, Denne M, Stallard P. Supported Web-Based Guided Self-Help for Insomnia for Young People Attending Child and Adolescent Mental Health Services: Protocol for a Feasibility Assessment. JMIR Res Protoc. 2018;7(12). doi:10.2196/11324

38. Robinson E, Titov N, Andrews G, McIntyre K, Schwencke G, Solley K. Internet treatment for generlized anxiety disorder: A randomized controlled trial comparing clinician vs. technician assistance. PLoS One. 2010;5(6). doi:10.1371/journal.pone.0010942

39. Titov N, Andrews G, Robinson E, et al. Clinician-assisted Internet-based treatment is effective for generalized anxiety disorder: randomized controlled trial. Australian & New Zealand Journal of Psychiatry. 2009;43(10):905–912. http://www.virtualclinic.org.au,

40. Titov N, Dear BF, Johnston L, et al. Improving Adherence and Clinical Outcomes in Self-Guided Internet Treatment for Anxiety and Depression: Randomised Controlled Trial. PLoS One. 2013;8(7). doi:10.1371/journal.pone.0062873

41. Rothmann MD, Wiens BL, Chan ISF. Design and Analysis of Non-Inferiority Trials. Chapman and Hall/CRC; 2016. doi:10.1201/b11039.

42. Catarino A, Bateup S, Tablan V, et al. Demographic and clinical predictors of response to internet-enabled cognitive–behavioural therapy for depression and anxiety. BJPsych Open. 2018;4(5):411–418. doi:10.1192/bjo.2018.57

43. Ho DE, Imai K, King G, Stuart EA. MatchIt: Nonparametric Preprocessing for Parametric Causal Inference. Vol 42.; 2011. http://www.jstatsoft.org/

44. R Core Team. R: A Language and Environment for Statistical Computing. R Foundation for Statistical Computing; 2016.

45. Toussaint A, Hüsing P, Gumz A, et al. Sensitivity to change and minimal clinically important difference of the 7-item Generalized Anxiety Disorder Questionnaire (GAD-7). J Affect Disord. 2020;265:395–401. doi:10.1016/j.jad.2020.01.032

46. Clark DM. Realizing the Mass Public Benefit of Evidence-Based Psychological Therapies: The IAPT Program. Annu Rev Clin Psychol. 2018;14:159–183. doi:10.1146/annurev-clinpsy-050817-084833

47. Ali S, Rhodes L, Moreea O, et al. How durable is the effect of low intensity CBT for depression and anxiety? Remission and relapse in a longitudinal cohort study. Behaviour Research and Therapy. 2017;94:1–8. doi:10.1016/j.brat.2017.04.006

48. Delgadillo J, Rhodes L, Moreea O, et al. Relapse and Recurrence of Common Mental Health Problems after Low Intensity Cognitive Behavioural Therapy: The WYLOW Longitudinal Cohort Study. Psychother Psychosom. 2018;87(2):116–117. doi:10.1159/000485386

49. Shallcross AJ, Willroth Aaron Fisher EC, Dimidjian S, Gross JJ, Visvanathan Manhattan Mindfulness-Based Cognitive Behavioral Therapy Iris B Mauss PD. Relapse/Recurrence Prevention in Major Depressive Disorder: 26-Month Follow-Up of Mindfulness-Based Cognitive Therapy Versus an Active Control ScienceDirect. Behav Ther. Published online 2018. http://www.sciencedirect.comwww.elsevier.com/locate/bt

50. NHS Digital. NHS Talking Therapies, for anxiety and depression, Annual reports, 2022-23. NHS Digital. January 12, 2024. Accessed May 24, 2024. https://digital.nhs.uk/data-and-information/publications/statistical/nhs-talking-therapies-for-anxiety-and-depression-annual-reports/2022-23#resources

51. Richards D, Enrique A, Eilert N, et al. A pragmatic randomized waitlist-controlled effectiveness and cost-effectiveness trial of digital interventions for depression and anxiety. NPJ Digit Med. 2020;3(1). doi:10.1038/s41746-020-0293-8

52. Jabir AI, Lin X, Martinengo L, Sharp G, Theng YL, Car LT. Attrition in Conversational Agent-Delivered Mental Health Interventions: Systematic Review and Meta-Analysis. J Med Internet Res. 2024;26(1). doi:10.2196/48168

53. Olfson M, Marcus S. National Trends in Outpatient Psychotherapy. Am J Psychiatry . 2010;167(12):1456–1463.

54. Wells JE, Browne MO, Aguilar-Gaxiola S, et al. Drop out from out-patient mental healthcare in the World Health Organization’s World Mental Health Survey initiative. British Journal of Psychiatry. 2013;202(1):42–49. doi:10.1192/bjp.bp.112.113134

55. U.S. Department of Health and Human Services Food and Drug Administration. Considerations for the Design and Conduct of Externally Controlled Trials for Drug and Biological Products Guidance for Industry DRAFT GUIDANCE.; 2023. https://www.fda.gov/vaccines-blood-biologics/guidance-compliance-regulatory-information-biologics/biologics-guidances

56. National Institute for Health and Care Excellence. NICE Real-World Evidence Framework.; 2022. http://www.nice.org.uk/corporate/ecd9

57. Thorlund K, Dron L, Park JJH, Mills EJ. Synthetic and external controls in clinical trials – A primer for researchers. Clin Epidemiol. 2020;12:457–467. doi:10.2147/CLEP.S242097

58. Corrigan-Curay J, Sacks L, Woodcock J. Real-world evidence and real-world data for evaluating drug safety and effectiveness. JAMA - Journal of the American Medical Association. 2018;320(9):867–868. doi:10.1001/jama.2018.10136

59. Patterson B, Boyle MH, Kivlenieks M, Van Ameringen M. The use of waitlists as control conditions in anxiety disorders research. J Psychiatr Res. 2016;83:112–120. doi:10.1016/j.jpsychires.2016.08.015

60. Thorlund K, Dron L, Park JJH, Mills EJ. Synthetic and external controls in clinical trials – A primer for researchers. Clin Epidemiol. 2020;12:457–467. doi:10.2147/CLEP.S242097

61. American Psychological Association. DIAGNOSIS: Mixed Anxiety Conditions TREATMENT: Acceptance And Commitment Therapy For Mixed Anxiety Disorders. 2015. Accessed June 24, 2024. https://div12.org/treatment/acceptance-and-commitment-therapy-for-mixed-anxiety-disorders/

62. Han A, Kim TH. Efficacy of Internet-Based Acceptance and Commitment Therapy for Depressive Symptoms, Anxiety, Stress, Psychological Distress, and Quality of Life: Systematic Review and Meta-analysis. J Med Internet Res. 2022;24(12). doi:10.2196/39727

63. Papola D, Miguel C, Mazzaglia M, et al. Psychotherapies for Generalized Anxiety Disorder in Adults: A Systematic Review and Network Meta-Analysis of Randomized Clinical Trials. JAMA Psychiatry. 2024;81(3):250–259. doi:10.1001/jamapsychiatry.2023.3971

64. Kelson J, Rollin A, Ridout B, Campbell A. Internet-delivered acceptance and commitment therapy for anxiety treatment: Systematic review. J Med Internet Res. 2019;21(1). doi:10.2196/12530

65. Hemmings NR, Kawadler JM, Whatmough R, et al. Development and feasibility of a digital acceptance and commitment therapy?based intervention for generalized anxiety disorder: Pilot acceptability study. JMIR Form Res. 2021;5(2). doi:10.2196/21737

66. Coghlan S, Leins K, Sheldrick S, Cheong M, Gooding P, D’Alfonso S. To chat or bot to chat: Ethical issues with using chatbots in mental health. Digit Health. 2023;9. doi:10.1177/20552076231183542

67. Huang YS (Sandy), Dootson P. Chatbots and service failure: When does it lead to customer aggression. Journal of Retailing and Consumer Services. 2022;68. doi:10.1016/j.jretconser.2022.103044

68. Nuffield Council on Bioethics. The Role of Technology in Mental Healthcare.; 2022. Accessed June 24, 2024. https://www.nuffieldbioethics.org/assets/pdfs/The-role-of-technology-in-mental-healthcare.pdf

69. Stade EC, Stirman SW, Ungar LH, et al. Large language models could change the future of behavioral healthcare: a proposal for responsible development and evaluation. npj Mental Health Research. 2024;3(1). doi:10.1038/s44184-024-00056-z

70. Flückiger C, Wampold BE, Delgadillo J, Rubel J, Vîsla A, Lutz W. Is There an Evidence-Based Number of Sessions in Outpatient Psychotherapy? - A Comparison of Naturalistic Conditions across Countries. Psychother Psychosom. 2020;89(5):333–335. doi:10.1159/000507793

71. Terlizzi E, Villarroel M. Symptoms of Generalized Anxiety Disorder Among Adults: United States, 2019.; 2020. Accessed July 4, 2024. https://www.cdc.gov/nchs/products/databriefs/db378.htm

