## Supplementary Information for "Combining AI and human support in mental health: a digital intervention with comparable effectiveness to human-delivered care"

#### Table of Contents


| Supplementary Methods | |
|---|---|
| A priori definition of non-inferiority margin | 2 |
| **Supplementary Tables** | |
| Supplementary Table 1. Description of each module topic in the digital program. | 3 |
| Supplementary Table 2. Propensity-matching between groups. | 4 |
| Supplementary Table 3. Output from slope analysis comparing adherence rate over GAD-7 check-ins across groups. | 5 |
| Supplementary Table 4. Output of linear regression model measuring the association between participant characteristics and adherence for the ITT sample. | 6 |
| Supplementary Table 5. Output of logistic regression model measuring association between participant characteristics and non-adherence. | 7 |
| Supplementary Table 6. Rates of improvement, recovery, reliable recovery, responder and remission from baseline to final score across all groups for the per-protocol sample. | 8 |
| Supplementary Table 7. Rates of improvement, recovery, reliable recovery, responder and remission from baseline to final score across all groups for the intention-to-treat sample | 9 |
| Supplementary Table 8. Mean GAD-7 score across assessments stratified by baseline GAD-7 severity. | 10 |
| Supplementary Table 9. Change in GAD-7, PHQ-9 and WSAS scores from baseline to final score stratified by baseline GAD-7 severity for the intervention sample. | 11 |
| Supplementary Table 10. Output of linear regression model measuring association between participant characteristics and change in GAD-7 scores for the ITT intervention sample. | 12 |
| Supplementary Table 11. Mean PHQ-9 score across assessments stratified by baseline GAD-7 severity. | 13 |

### Supplementary Methods

*A priori definition of non-inferiority margin*

The Food and Drugs Administration (FDA) recommends that the threshold for non-inferiority is set based on estimates of the active comparator in previously conducted studies (70). This threshold can be defined, for example, as 50% or less of the lower confidence interval of the expected effect of the active comparator vs placebo. While it is difficult to develop an appropriate placebo in the context of psychotherapy research, previous studies exploring the efficacy of internet-delivered psychotherapy for GAD, against a waiting list control group, demonstrate significant clinical benefits of these interventions. In these studies, medium to large between-group effect sizes are reported, ranging from 0.38 to 1.25 (38–40). The outcomes from these studies are aggregated using a fixed effect meta-analysis, results shown below.

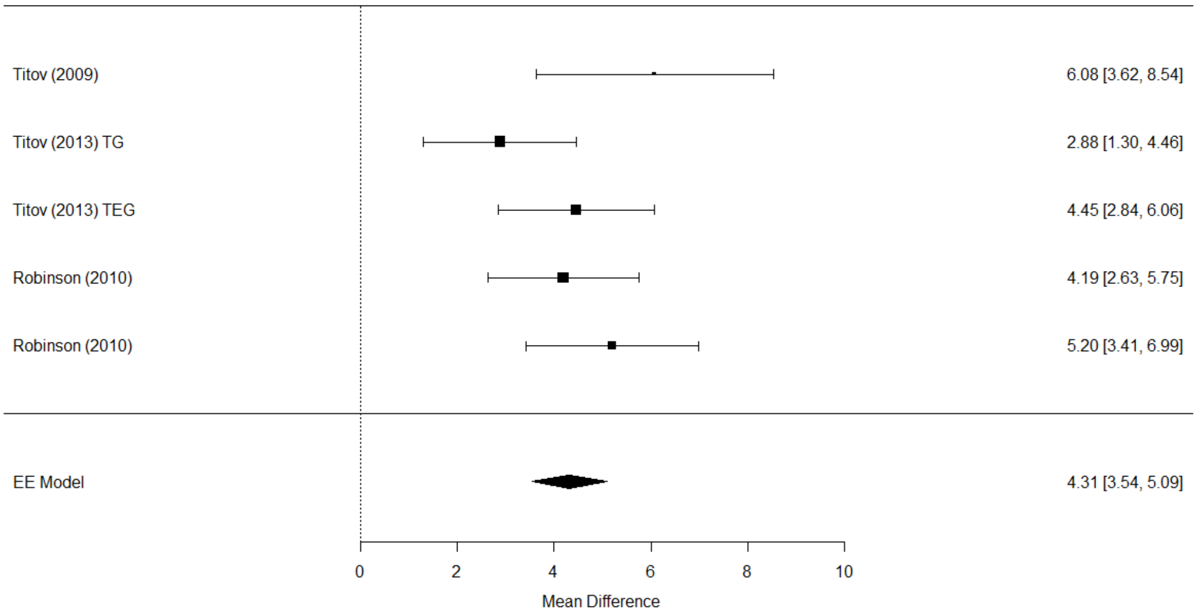


Here we define the non-inferiority margin as a GAD-7 score change of 1.8. This is equivalent to 50% of the lower limit of the 95% confidence interval of the fixed effect meta-analysis pooled result (i.e. 3.54/2).

### Supplementary Tables

**Supplementary Table 1. Description of each module topic in the digital program.** The digital program consisted of six modules with three sessions each following a pattern of (1) learning; (2) activity; (3) practice. The final modules (5 + 6) focused on consolidation of learning.

| Module | Topic | Description |
|---|---|---|
| 1 | Getting to Know You + Learning About Worry & Anxiety | Questions, reflections and information designed to help increase understanding of difficulties and prepare for making change. |
| 2 | Holding Thoughts Lightly | Focus on unhelpful patterns of thinking with activities and practices designed to promote more flexible ways to respond to thoughts. |
| 3 | Making Meaningful Moves Towards What Matters | Focus on understanding and targeting unhelpful avoidance behaviors, with activities and practices designed to gradually reduce these unhelpful behaviors. |
| 4 | Taking a Different Perspective | Focus on unhelpful beliefs about worry which may be maintaining anxiety symptoms, with activities and practices designed to reframe these beliefs. |
| 5 + 6 | Continuing On Your Journey and Bringing it All Together | Focus on consolidation of skills and techniques. Planning on maintaining progress once the program has been completed. |

**Supplementary Table 2. Demographic data for propensity-matched groups.** Demographic data used to propensity match across the intervention sample and external control groups: age, baseline anxiety symptoms (GAD-7), baseline mood symptoms (PHQ-9) and presence of a chronic health condition. Similarity across groups in means (continuous variables) and proportions (categorical variable) highlights success of propensity-matching.

| | Per-protocol | | Intention-to-treat | | |
|---|---|---|---|---|---|
| Variable | ieso Digital Program (N=169) | Waiting control (N=169) | ieso Digital Program (N=299) | Face-to-face CBT (N=299) | Typed CBT (N=299) |
| Age, Mean (SD) | 41.7 (11.8) | 41.7 (13.3) | 39.8 (12.8) | 40.1 (16.6) | 39.8 (12.7) |
| Baseline GAD-7, Mean (SD) | 12.4 (3.4) | 12.5 (3.3) | 12.5 (3.3) | 12.9 (3.1) | 12.6 (3.5) |
| Baseline PHQ-9, Mean (SD) | 8.0 (3.8) | 8.4 (3.4) | 8.0 (3.7) | 8.4 (3.6) | 8.1 (3.6) |
| Chronic health condition: Yes, N (%) | 70 (41.4) | 78 (46.2) | 114 (38.1) | 119 (39.8) | 108 (36.1) |
| Chronic health condition: No, N (%) | 91 (53.8) | 90 (53.3) | 167 (55.9) | 180 (60.2) | 173 (57.9) |
| Chronic health condition: Not Known, N (%) | 8 (4.7) | 1 (0.6) | 18 (6.0) | 0 (0.0) | 18 (6.0) |

**Supplementary Table 3. Comparison of adherence rates over sessions across groups.** For each group, adherence was defined based on the proportion of participants who completed each GAD-7 assessment ("symptom check") throughout their journey either within the ieso digital program or at the start of their treatment session for the control groups across seven total instances of GAD-7 data collection. A) Slopes were estimated using a regression model predicting adherence by session number to quantify the adherence rate for each group. B) To test for differences in the adherence rate across groups a generalized linear regression model was used. Non-significant session-by-group interaction indicates no significant difference between groups in adherence rate. Reference value = ieso digital program. * = $p < .05$; ** = $p < .005$

*A) Adherence rate estimates per group*

| Group | Slope | Std. Error | L95%CI | U95%CI |
|---|---|---|---|---|
| ieso Digital Program | −10.29 | 0.26 | −10.80 | −9.78 |
| Face-to-face CBT | −9.55 | 0.31 | −10.16 | −8.95 |
| Typed CBT | −10.99 | 0.58 | −12.14 | −9.85 |

*B) Generalized linear regression model output*

| Variable | Estimate | Std. Error | t value | p-value | significance |
|---|---|---|---|---|---|
| (Intercept) | 113.378 | 7.634 | 14.851 | 6.57e-14 | ** |
| Session | −10.870 | 1.707 | −6.367 | 1.15e-06 | ** |
| Group: Face-to-face CBT | −14.650 | 9.444 | −1.551 | 0.133 | |
| Group: Typed CBT | −8.518 | 9.444 | −0.902 | 0.376 | |
| Session*Group: Face-to-face CBT | 2.567 | 1.867 | 1.375 | 0.181 | |
| Session*Group: Typed CBT | 0.586 | 1.867 | 0.314 | 0.756 | |

**Supplementary Table 4. Linear regression model measuring the association between participant characteristics and adherence for the ITT sample.** Adherence was defined as the number of completed sessions in the digital program. All demographic data, GAD-7 score, and PHQ-9 score were collected at baseline. Enrolment path refers to whether a participant was a referred patient through NHS TT or was externally recruited. Reference values for each categorical variable were: chronic health condition = yes; gender = female; software version = 1; disability = yes; sexual orientation = heterosexual; employment = employed; ethnicity = white; qualification = degree; religion = Christian; enrolment path = NHS patient; Medication = Not taking medication. * = $p < .05$; ** = $p < .005$.

| Variable | Estimate | Std. Error | t value | p-value | significance |
|---|---|---|---|---|---|
| (Intercept) | 10.058 | 3.403 | 2.956 | 0.003 | ** |
| Baseline score: GAD-7 | −0.081 | 0.156 | −0.520 | 0.604 | |
| Baseline score :PHQ-9 | 0.060 | 0.134 | 0.446 | 0.656 | |
| Age (at screening) | 0.112 | 0.042 | 2.646 | 0.009 | ** |
| Chronic health condition: no | −0.155 | 1.093 | −0.142 | 0.887 | |
| Chronic health condition: not known | −2.142 | 2.576 | −0.832 | 0.406 | |
| Gender: male | −1.494 | 1.329 | −1.124 | 0.262 | |
| Gender: other | −1.197 | 4.248 | −0.282 | 0.778 | |
| Gender: not known | −0.714 | 5.169 | −0.138 | 0.890 | |
| Software version: version 2 | 0.097 | 1.172 | 0.083 | 0.934 | |
| Disability: no perceived disability | 0.627 | 1.301 | 0.482 | 0.630 | |
| Disability: prefer not to say | −1.545 | 3.907 | −0.395 | 0.693 | |
| Sexual orientation: not known | 2.059 | 2.797 | 0.736 | 0.462 | |
| Sexual orientation: other | 1.742 | 1.399 | 1.245 | 0.214 | |
| Employment: not employed | −1.921 | 1.375 | −1.397 | 0.163 | |
| Employment: not known | −6.595 | 3.405 | −1.937 | 0.054 | . |
| Ethnicity: not known | 4.388 | 5.130 | 0.855 | 0.393 | |
| Ethnicity: other | 0.750 | 1.872 | 0.400 | 0.689 | |
| Qualification: below degree | −1.373 | 1.216 | −1.129 | 0.260 | |
| Qualification: not known | −2.050 | 6.092 | −0.336 | 0.737 | |
| Qualification: other | −1.320 | 3.837 | −0.344 | 0.731 | |
| Qualification: postgraduate | 1.788 | 1.138 | 1.571 | 0.117 | |
| Religion: not known | −0.854 | 3.030 | −0.282 | 0.778 | |
| Religion: other | −1.057 | 1.139 | −0.928 | 0.354 | |
| Enrolment path: external recruit | 0.882 | 1.121 | 0.787 | 0.432 | |
| Medication: Taking medication | 0.052 | 1.027 | 0.051 | 0.959 | |

Residual standard error: 7.849 on 273 degrees of freedom
Multiple R-squared: 0.1091, Adjusted R-squared: 0.02756
F-statistic: 1.338 on 25 and 273 DF, p-value: 0.1347

**Supplementary Table 5. Logistic regression model measuring association between participant characteristics and non-adherence.** Non-adherence was defined based on PP sample status, i.e. non-adherence (coded = 1) included all those *not* in the PP sample. All demographic data, GAD-7 score and PHQ-9 score were collected at baseline. Enrolment path refers to whether a participant was a referred patient through NHS TT or was externally recruited. Reference values for each categorical variable were: chronic health condition = yes; gender = female; software version = 1; disability = yes; sexual orientation = heterosexual; employment = employed; ethnicity = white; qualification = degree; religion = Christian; enrolment path = NHS patient; Medication = Not taking medication. * = $p < .05$.

| Variable | Estimate | Odds Ratio | 95% LCI | 95% UCI | Std. Error | z value | p-value | Significance |
|---|---|---|---|---|---|---|---|---|
| (Intercept) | 0.877 | 2.404 | 0.398 | 14.526 | 0.918 | 0.956 | 0.339 | |
| Baseline score: GAD-7 | −0.001 | 0.999 | 0.920 | 1.085 | 0.042 | −0.026 | 0.979 | |
| Baseline score :PHQ-9 | −0.026 | 0.974 | 0.908 | 1.045 | 0.036 | −0.735 | 0.462 | |
| Age (at screening) | −0.029 | 0.971 | 0.949 | 0.993 | 0.012 | −2.543 | 0.011 | * |
| Chronic health condition: no | 0.225 | 1.253 | 0.701 | 2.238 | 0.296 | 0.761 | 0.447 | |
| Chronic health condition: not known Known | 0.249 | 1.282 | 0.338 | 4.862 | 0.680 | 0.366 | 0.715 | |
| Gender: male | 0.205 | 1.227 | 0.608 | 2.476 | 0.358 | 0.571 | 0.568 | |
| Gender: other | 0.241 | 1.273 | 0.138 | 11.731 | 1.133 | 0.213 | 0.831 | |
| Gender: not known | −0.350 | 0.705 | 0.027 | 18.102 | 1.656 | −0.211 | 0.833 | |
| Product version: version 2 | 0.005 | 1.005 | 0.542 | 1.867 | 0.316 | 0.017 | 0.986 | |
| Disability: no perceived disability | −0.197 | 0.822 | 0.411 | 1.642 | 0.353 | −0.557 | 0.578 | |
| Disability: prefer not to say | 0.737 | 2.090 | 0.249 | 17.572 | 1.086 | 0.679 | 0.497 | |
| Sexual orientation: not known | −0.548 | 0.578 | 0.129 | 2.598 | 0.767 | −0.715 | 0.475 | |
| Sexual orientation: other | −0.399 | 0.671 | 0.318 | 1.417 | 0.381 | −1.046 | 0.295 | |
| Employment: not employed | 0.693 | 2.000 | 0.964 | 4.151 | 0.372 | 1.861 | 0.063 | . |
| Employment: not known | 1.193 | 3.296 | 0.512 | 21.234 | 0.950 | 1.255 | 0.210 | |
| Ethnicity: not known | −0.647 | 0.524 | 0.020 | 13.633 | 1.663 | −0.389 | 0.697 | |
| Ethnicity: other | 0.147 | 1.158 | 0.430 | 3.117 | 0.505 | 0.290 | 0.772 | |
| Qualification: below degree | 0.495 | 1.641 | 0.865 | 3.114 | 0.327 | 1.516 | 0.130 | |
| Qualification: not known | 0.466 | 1.593 | 0.070 | 36.143 | 1.593 | 0.292 | 0.770 | |
| Qualification: other | 1.078 | 2.938 | 0.324 | 26.654 | 1.125 | 0.958 | 0.338 | |
| Qualification: postgraduate | −0.076 | 0.927 | 0.505 | 1.700 | 0.309 | −0.245 | 0.806 | |
| Religion: not known | −0.192 | 0.826 | 0.162 | 4.204 | 0.830 | −0.231 | 0.817 | |
| Religion: other | 0.405 | 1.499 | 0.814 | 2.760 | 0.311 | 1.299 | 0.194 | |
| Enrolment path: external recruit | −0.381 | 0.683 | 0.381 | 1.224 | 0.298 | −1.281 | 0.200 | |
| Medication: Taking medication | −0.065 | 0.937 | 0.544 | 1.614 | 0.277 | −0.235 | 0.814 | |

Null deviance: 409.40 on 298 degrees of freedom
Residual deviance: 380.45 on 273 degrees of freedom; p-value = 1.796641e-05
AIC: 432.45

**Supplementary Table 6. Rates of improvement, recovery, reliable recovery, responder and remission from baseline to final score across all groups for the per-protocol sample.** Chi-squared tests were conducted to test for differences in binary clinical outcomes between the ieso digital program and each external control group. Significant tests after Bonferroni correction for multiple comparisons based on number of outcome metrics (n=7) in bold.

| | N improved | N not improved | N total | % improved | % not improved | $\chi^2$ | *p* | *corr-p* |
|---|---|---|---|---|---|---|---|---|
| *ieso Digital Program* | 139 | 30 | 169 | 82.2 | 17.8 | | | |
| *Waiting control* | 61 | 108 | 169 | 36.1 | 63.9 | **72.6** | **<.001** | **<.001** |
| *Face-to-face CBT* | 189 | 64 | 253 | 74.7 | 25.3 | 2.9 | 0.088 | .616 |
| *Typed CBT* | 195 | 34 | 229 | 85.2 | 14.8 | 0.4 | 0.521 | 1.000 |
| | N recovered | N not recovered | N total | % recovered | % not recovered | $\chi^2$ | *p* | *corr-p* |
| *ieso Digital Program* | 130 | 39 | 169 | 76.9 | 23.1 | | | |
| *Waiting control* | 46 | 123 | 169 | 27.2 | 72.8 | **81.7** | **<.001** | **<.001** |
| *Face-to-face CBT* | 164 | 89 | 253 | 64.8 | 35.2 | 6.5 | 0.011 | .077 |
| *Typed CBT* | 196 | 33 | 229 | 85.6 | 14.4 | 4.4 | 0.037 | .259 |
| | N reliable recovery | N not reliable recovery | N total | % reliable recovery | % not reliable recovery | $\chi^2$ | *p* | *corr-p* |
| *ieso Digital Program* | 122 | 47 | 169 | 72.2 | 27.8 | | | |
| *Waiting control* | 34 | 135 | 169 | 20.1 | 79.9 | **90.1** | **<.001** | **<.001** |
| *Face-to-face CBT* | 157 | 96 | 253 | 62.1 | 37.9 | 4.2 | 0.04 | .280 |
| *Typed CBT* | 180 | 49 | 229 | 78.6 | 21.4 | 1.8 | 0.174 | 1.000 |
| | N GAD remission | N not GAD remission | N total | % GAD remission | % not GAD remission | $\chi^2$ | *p* | *corr-p* |
| *ieso Digital Program* | 136 | 33 | 169 | 80.5 | 19.5 | | | |
| *Waiting control* | 48 | 121 | 169 | 28.4 | 71.6 | **90.3** | **<.001** | **<.001** |
| *Face-to-face CBT* | 170 | 83 | 253 | 67.2 | 32.8 | **8.3** | **0.004** | **.028** |
| *Typed CBT* | 198 | 31 | 229 | 86.5 | 13.5 | 2.2 | 0.142 | .994 |
| | N GAD responder | N not GAD responder | N total | % GAD responder | % not GAD responder | $\chi^2$ | *p* | *corr-p* |
| *ieso Digital Program* | 138 | 31 | 169 | 81.7 | 18.3 | | | |
| *Waiting control* | 56 | 113 | 169 | 33.1 | 66.9 | **79.4** | **<.001** | **<.001** |
| *Face-to-face CBT* | 185 | 68 | 253 | 73.1 | 26.9 | 3.6 | 0.056 | .392 |
| *Typed CBT* | 192 | 37 | 229 | 83.8 | 16.2 | 0.2 | 0.661 | 1.000 |
| | N PHQ remission | N not PHQ remission | N total | % PHQ remission | % not PHQ remission | $\chi^2$ | *p* | *corr-p* |
| *ieso Digital Program* | 61 | 8 | 69 | 88.4 | 11.6 | | | |
| *Waiting control* | 36 | 30 | 66 | 54.5 | 45.5 | **17.5** | **<.001** | **<.001** |
| *Face-to-face CBT* | 84 | 19 | 103 | 81.6 | 18.4 | 1 | 0.319 | 1.000 |
| *Typed CBT* | 66 | 10 | 76 | 86.8 | 13.2 | 0.001 | 0.974 | 1.000 |
| | N PHQ responder | N not PHQ responder | N total | %PHQ responder | % not PHQ responder | $\chi^2$ | *p* | *corr-p* |
| *ieso Digital Program* | 46 | 123 | 169 | 27.2 | 72.8 | | | |
| *Waiting control* | 16 | 153 | 169 | 9.5 | 90.5 | **16.6** | **<.001** | **<.001** |
| *Face-to-face CBT* | 78 | 175 | 253 | 30.8 | 69.2 | 0.5 | 0.491 | 1.000 |
| *Typed CBT* | 81 | 148 | 229 | 35.4 | 64.6 | 2.6 | 0.106 | .742 |

**Supplementary Table 7. Rates of improvement, recovery, reliable recovery, responder and remission from baseline to final score across all groups for the intention-to-treat sample.** Chi-squared tests were conducted to test for differences in binary clinical outcomes between the ieso digital program and each external control group. Significant tests after Bonferroni correction for multiple comparisons based on number of outcome metrics (n=7) in bold.

| | N improved | N not improved | N total | % improved | % not improved | $\chi^2$ | *p* | *corr-p* |
|---|---|---|---|---|---|---|---|---|
| *ieso Digital Program* | 198 | 101 | 299 | 66.2 | 33.8 | | | |
| *Face-to-face CBT* | 211 | 88 | 299 | 70.6 | 29.4 | 1.1 | 0.291 | 1.000 |
| *Typed CBT* | 230 | 69 | 299 | 76.9 | 23.1 | **7.9** | **0.005** | **.035** |
| | N recovered | N not recovered | N total | % recovered | % not recovered | $\chi^2$ | *p* | *corr-p* |
| *ieso Digital Program* | 174 | 125 | 299 | 58.2 | 41.8 | | | |
| *Face-to-face CBT* | 189 | 110 | 299 | 63.2 | 36.8 | 1.4 | 0.241 | 1.000 |
| *Typed CBT* | 226 | 73 | 299 | 75.6 | 24.4 | **19.6** | **<.001** | **<.001** |
| | N reliable recovery | N not reliable recovery | N total | % reliable recovery | % not reliable recovery | $\chi^2$ | *p* | *corr-p* |
| *ieso Digital Program* | 161 | 138 | 299 | 53.8 | 46.2 | | | |
| *Face-to-face CBT* | 178 | 121 | 299 | 59.5 | 40.5 | 1.7 | 0.187 | 1.000 |
| *Typed CBT* | 204 | 95 | 299 | 68.2 | 31.8 | **12.4** | **<.001** | **.003** |
| | N GAD remission | N not GAD remission | N total | % GAD remission | % not GAD remission | $\chi^2$ | *p* | *corr-p* |
| *ieso Digital Program* | 184 | 115 | 299 | 61.5 | 38.5 | | | |
| *Face-to-face CBT* | 196 | 103 | 299 | 65.6 | 34.4 | 0.9 | 0.35 | 1.000 |
| *Typed CBT* | 229 | 70 | 299 | 76.6 | 23.4 | **15.2** | **<.001** | **.001** |
| | N GAD responder | N not GAD responder | N total | % GAD responder | % not GAD responder | $\chi^2$ | *p* | *corr-p* |
| *ieso Digital Program* | 199 | 100 | 299 | 66.6 | 33.4 | | | |
| *Face-to-face CBT* | 207 | 92 | 299 | 69.2 | 30.8 | 0.4 | 0.54 | 1.000 |
| *Typed CBT* | 228 | 71 | 299 | 76.3 | 23.7 | 6.4 | 0.011 | .077 |
| | N PHQ remission | N not PHQ remission | N total | % PHQ remission | % not PHQ remission | $\chi^2$ | *p* | *corr-p* |
| *ieso Digital Program* | 78 | 38 | 116 | 67.2 | 12.7 | | | |
| *Face-to-face CBT* | 89 | 27 | 116 | 76.7 | 9.0 | 2.1 | 0.144 | 1.000 |
| *Typed CBT* | 82 | 22 | 104 | 78.8 | 7.4 | 3.2 | 0.075 | .525 |
| | N PHQ responder | N not PHQ responder | N total | %PHQ responder | % not PHQ responder | $\chi^2$ | *p* | *corr-p* |
| *ieso Digital Program* | 55 | 244 | 299 | 18.4 | 81.6 | | | |
| *Face-to-face CBT* | 86 | 213 | 299 | 28.8 | 71.2 | **8.4** | **0.004** | **.028** |
| *Typed CBT* | 88 | 211 | 299 | 29.4 | 70.6 | **9.4** | **0.002** | **.014** |

**Supplementary Table 8. Mean GAD-7 score across assessments stratified by baseline GAD-7 severity.** Mean GAD-7 scores for both PP and ITT samples. Stratification by severity based on baseline GAD-7 score. Check-in scores were collected prior to each module within the digital program software. Screening, completion and follow-up scores were collected outside of the digital program. Data reported are for each specific data collection point.

| Study timepoint | Sample | Per-protocol | | | | Intention-to-treat | | | |
|---|---|---|---|---|---|---|---|---|---|
| | Severity | Mild | Moderate | Severe | Overall | Mild | Moderate | Severe | Overall |
| Screening | N | 39 | 82 | 48 | 169 | 62 | 150 | 87 | 299 |
| | Mean (95%CIs) | 8.6 (8.4, 8.7) | 11.7 (11.3, 12.0) | 16.9 (16.4, 17.4) | 12.4 (11.9, 13.0) | 8.5 (8.4, 8.6) | 11.7 (11,5, 11.9) | 16.9 (16.5, 17.2) | 12.5 (12.2, 12.9) |
| Check-in 1 | N | 39 | 82 | 48 | 169 | 59 | 144 | 81 | 284 |
| | Mean (95%CIs) | 9.8 (8.5, 11.1) | 11.2 (10.4, 12.0) | 14.0 (12.9, 15.2) | 11.7 (11.1, 12.3) | 9.9 (8.9, 10.9) | 10.9 (10.3, 11.5) | 14.0 (13.2, 14.9) | 11.6 (11.1, 12.1) |
| Check-in 2 | N | 39 | 82 | 48 | 169 | 54 | 120 | 66 | 240 |
| | Mean (95%CIs) | 6.0 (5.1, 6.9) | 8.1 (7.3, 8.9) | 9.2 (8.1, 10.4) | 7.9 (7.4, 8.5) | 6.4 (5.5, 7.3) | 8.1 (7.4, 8.7) | 9.6 (8.5, 10.7) | 8.1 (7.6, 8.6) |
| Check-in 3 | N | 39 | 82 | 48 | 169 | 46 | 106 | 57 | 209 |
| | Mean (95%CIs) | 5.8 (4.7, 6.8) | 7.0 (6.2, 7.9) | 8.2 (6.9, 9.5) | 7.1 (6.5, 7.7) | 5.6 (4.7, 6.6) | 7.2 (6.5, 8.0) | 8.5 (7.3, 9.7) | 7.2 (6.7, 7.8) |
| Check-in 4 | N | 39 | 82 | 48 | 169 | 40 | 90 | 50 | 180 |
| | Mean (95%CIs) | 4.5 (3.7, 5.3) | 5.9 (5.1, 6.6) | 7.0 (5.8, 8.2) | 5.8 (5.3, 6.4) | 4.8 (3.8, 5.7) | 5.9 (5.2, 6.6) | 7.1 (5.9, 8.3) | 6.0 (5.5, 6.5) |
| Check-in 5 | N | 31 | 68 | 36 | 135 | 31 | 70 | 37 | 138 |
| | Mean (95%CIs) | 4.7 (3.6, 5.8) | 5.4 (4.6, 6.1) | 6.9 (5.2, 8.6) | 5.6 (5.0, 6.2) | 4.7 (3.6, 5.8) | 5.3 (4.6, 6.0) | 7.3 (5.5, 9.1) | 5.7 (5.0, 6.3) |
| Check-in 6 | N | 23 | 61 | 28 | 112 | 23 | 62 | 28 | 113 |
| | Mean (95%CIs) | 3.9 (2.8, 5.1) | 5.2 (4.4, 6.1) | 6.5 (4.6, 8.4) | 5.3 (4.6, 6.0) | 3.9 (2.8, 5.1) | 5.2 (4.3, 6.0) | 6.5 (4.6, 8.4) | 5.2 (4.5, 5.9) |
| Completion | N | 39 | 82 | 48 | 169 | 46 | 101 | 56 | 203 |
| | Mean (95%CIs) | 4.2 (3.3, 5.1) | 4.8 (4.2, 5.5) | 6.2 (4.8, 7.6) | 5.1 (4.5, 5.6) | 4.3 (3.4, 5.1) | 5.4 (4.7, 6.0) | 6.7 (5.3, 8.0) | 5.5 (4.9, 6.0) |
| Follow-up | N | 39 | 80 | 47 | 166 | 48 | 106 | 56 | 210 |
| | Mean (95%CIs) | 4.5 (3.5, 5.4) | 4.7 (4.0, 5.5) | 6.3 (4.8, 7.6) | 5.1 (4.5, 5.7) | 4.6 (3.8 ; 5.5) | 5.5 (4.7, 6.2) | 6.7 (5.3, 8.1) | 5.6 (5.0, 6.2) |

**Supplementary Table 9. Change in GAD-7, PHQ-9 and WSAS scores from baseline to final score stratified by baseline GAD-7 severity for the intervention sample.** Change (i.e. mean difference) in GAD-7, PHQ-9 and WSAS scores was calculated using the difference between baseline and final scores for the digital intervention group. A negative mean difference denotes a reduction in scores. Within-subject effect sizes (Cohen's d) calculated for the total sample and each severity subgroup based on baseline GAD-7 severity stratification (shown as absolute values).

| Sample | GAD-7 severity | N | Baseline score | | Change in score | | | | |
|---|---|---|---|---|---|---|---|---|---|
| | | | Mean | SD | Mean | SD | Lower 95% CI | Upper 95% CI | Within-subjects effect size (*d*) |
| *GAD-7* | | | | | | | | | |
| Per-protocol | Mild | 39 | 8.6 | 0.5 | −4.4 | 3.0 | −5.3 | −3.4 | 1.4 |
| | Moderate | 82 | 11.7 | 1.5 | −6.8 | 3.4 | −7.6 | −6.1 | 2.0 |
| | Severe | 48 | 16.9 | 1.7 | −10.7 | 5.3 | −12.3 | −9.2 | 2.0 |
| | **Overall** | 169 | 12.4 | 3.4 | −7.4 | 4.6 | −8.1 | −6.7 | 1.6 |
| Intention-to-treat | Mild | 62 | 8.5 | 0.5 | −2.9 | 4.1 | −4.0 | −1.9 | 0.7 |
| | Moderate | 150 | 11.7 | 1.4 | −5.0 | 4.2 | −5.7 | −4.3 | 1.2 |
| | Severe | 87 | 16.9 | 1.6 | −7.9 | 6.0 | −9.2 | −6.6 | 1.3 |
| | **Overall** | 299 | 12.5 | 3.3 | −5.4 | 5.1 | −6.0 | −4.8 | 1.1 |
| *PHQ-9* | | | | | | | | | |
| Per-protocol | Mild | 39 | 6.9 | 3.5 | −2.3 | 3.0 | −3.3 | −1.3 | 0.8 |
| | Moderate | 82 | 7.5 | 3.7 | −2.7 | 4.4 | −3.7 | −1.8 | 0.6 |
| | Severe | 48 | 9.7 | 3.8 | −4.5 | 5.5 | −6.1 | −2.9 | 0.8 |
| | **Overall** | 169 | 8.0 | 3.8 | −3.1 | 4.5 | −3.8 | −2.4 | 0.7 |
| Intention-to-treat | Mild | 62 | 6.7 | 3.1 | −1.3 | 4.0 | −2.3 | −0.3 | 0.3 |
| | Moderate | 150 | 7.5 | 3.5 | −1.3 | 4.6 | −2.0 | −0.6 | 0.3 |
| | Severe | 87 | 9.7 | 3.9 | −2.3 | 5.5 | −3.5 | −1.2 | 0.4 |
| | **Overall** | 299 | 8.0 | 3.7 | −1.6 | 4.8 | −2.1 | −1.1 | 0.3 |
| *WSAS* | | | | | | | | | |
| Per-protocol | Mild | 39 | 12.9 | 5.9 | −3.7 | 6.0 | −5.6 | −1.7 | 0.6 |
| | Moderate | 82 | 15.1 | 6.7 | −5.7 | 6.0 | −7.0 | −4.3 | 0.9 |
| | Severe | 48 | 17.4 | 5.5 | −6.0 | 6.4 | −7.9 | −4.2 | 0.9 |
| | **Overall** | 169 | 15.3 | 6.4 | −5.3 | 6.2 | −6.2 | −4.4 | 0.9 |
| Intention-to-treat | Mild | 58 | 12.2 | 5.6 | −3.4 | 6.0 | −5.2 | −1.6 | 0.6 |
| | Moderate | 150 | 14.9 | 6.9 | −4.8 | 6.4 | −6.1 | −3.5 | 0.8 |
| | Severe | 87 | 16.8 | 6.1 | −5.5 | 6.9 | −7.4 | −3.7 | 0.8 |
| | **Overall** | 295 | 14.9 | 6.6 | −4.7 | 6.5 | −5.6 | −3.8 | 0.7 |

**Supplementary Table 10. Output of linear regression model measuring association between participant characteristics and change in GAD-7 scores for the ITT intervention sample.** Dependent variable was change in GAD-7 score from baseline to final score. All demographic data, GAD-7 score and PHQ-9 score were collected at baseline. Enrolment path refers to whether a participant was a referred patient from NHS TT or was externally recruited. Reference values for each categorical variable were: chronic health condition = yes; gender = female; software version = 1; disability = yes; sexual orientation = heterosexual; employment = employed; ethnicity = white; qualification = degree; religion = Christian; enrolment path = NHS patient; Medication = Not taking medication. * = $p < .05$; ** = $p < .005$; *** = $p < .001$

| Variable | Estimate | Std. Error | t value | p-value | significance |
|---|---|---|---|---|---|
| (Intercept) | −4.016 | 2.017 | −1.991 | 0.048 | * |
| Baseline score: GAD-7 | 0.692 | 0.093 | 7.458 | 0.000 | *** |
| Baseline score: PHQ-9 | −0.125 | 0.079 | −1.573 | 0.117 | |
| Age (at screening) | 0.076 | 0.025 | 3.016 | 0.003 | ** |
| Chronic health condition: no | −0.519 | 0.648 | −0.801 | 0.424 | |
| Chronic health condition: not known | −1.756 | 1.527 | −1.150 | 0.251 | |
| Gender: male | 0.178 | 0.788 | 0.226 | 0.822 | |
| Gender: other | −2.493 | 2.519 | −0.990 | 0.323 | |
| Gender: not known | 0.236 | 3.065 | 0.077 | 0.939 | |
| Product version: version 2 | 0.012 | 0.695 | 0.017 | 0.986 | |
| Disability: no perceived disability | −0.251 | 0.771 | −0.325 | 0.745 | |
| Disability: prefer not to say | −0.174 | 2.316 | −0.075 | 0.940 | |
| Sexual orientation: not known | −1.132 | 1.658 | −0.682 | 0.496 | |
| Sexual orientation: other | 0.811 | 0.829 | 0.978 | 0.329 | |
| Employment: not employed | −1.589 | 0.815 | −1.950 | 0.052 | . |
| Employment: not known | −1.083 | 2.019 | −0.537 | 0.592 | |
| Ethnicity: not known | 1.026 | 3.041 | 0.337 | 0.736 | |
| Ethnicity: other | 1.198 | 1.110 | 1.079 | 0.281 | |
| Qualification: below degree | −0.364 | 0.721 | −0.504 | 0.614 | |
| Qualification: not known | −1.231 | 3.612 | −0.341 | 0.734 | |
| Qualification: other | −0.378 | 2.275 | −0.166 | 0.868 | |
| Qualification: postgraduate | −0.764 | 0.675 | −1.132 | 0.259 | |
| Religion: not known | 0.043 | 1.797 | 0.024 | 0.981 | |
| Religion: other | 0.071 | 0.675 | 0.106 | 0.916 | |
| Enrolment path: external recruit | −0.228 | 0.664 | −0.343 | 0.732 | |
| Medication: Taking medication | −0.227 | 0.609 | −0.373 | 0.709 | |

Residual standard error: 4.653 on 273 degrees of freedom
Multiple R-squared: 0.232, Adjusted R-squared: 0.1621
F-statistic: 3.306 on 25 and 273 DF, p-value: 6.435e-07

**Supplementary Table 11. Mean PHQ-9 score across assessments stratified by baseline GAD-7 severity.** Mean scores for both PP and ITT samples. Stratification by severity based on baseline GAD-7 scores. Check-in scores were collected prior to each module within the digital program software. Screening, completion and follow-up scores were collected outside of the digital program. Data reported are for each specific data collection point.

| Study timepoint | Sample | Per-protocol | | | | Intention-to-treat | | | |
|---|---|---|---|---|---|---|---|---|---|
| | Severity | Mild | Moderate | Severe | Overall | Mild | Moderate | Severe | Overall |
| Screening | N | 39 | 82 | 48 | 169 | 62 | 150 | 87 | 299 |
| | Mean (95%CIs) | 6.9 (5.7, 8.0) | 7.5 (6.7; 8.3) | 9.7 (8.6; 10.8) | 8.0 (7.4; 8.6) | 6.7 (5.9, 7.5) | 7.5 (6.9, 8.1) | 9.7 (8.9, 10.5) | 8.0 (7.5, 8.4) |
| Check-in 1 | N | 39 | 82 | 48 | 169 | 59 | 144 | 81 | 284 |
| | Mean (95%CIs) | 9.7 (8.1, 11.3) | 10.6 (9.7, 11.5) | 12.5 (11.3, 13.8) | 10.9 (10.2, 11.6) | 9.2 (8.0, 10.4) | 10.3 (9.6, 10.9) | 12.3 (11.3, 13.3) | 10.6 (10.2, 11.2) |
| Check-in 2 | N | 39 | 82 | 48 | 169 | 54 | 120 | 66 | 240 |
| | Mean (95%CIs) | 6.7 (5.4, 7.9) | 7.5 (6.5, 8.4) | 7.7 (6.5, 8.9) | 7.3 (6.7, 8.0) | 6.5 (5.5, 7.6) | 7.3 (6.6, 8.0) | 8.0 (6.9, 9.1) | 7.3 (6.8, 7.9) |
| Check-in 3 | N | 39 | 82 | 48 | 169 | 46 | 106 | 57 | 209 |
| | Mean (95%CIs) | 5.7 (4.7, 6.7) | 6.5 (5.5, 7.4) | 6.9 (5.7, 8.2) | 6.4 (5.8, 7.0) | 5.6 (4.6, 6.5) | 6.4 (5.6, 7.2) | 6.9 (5.8, 8.1) | 6.4 (5.8, 6.9) |
| Check-in 4 | N | 39 | 82 | 48 | 169 | 40 | 90 | 50 | 180 |
| | Mean (95%CIs) | 5.1 (4.0, 6.2) | 5.9 (5.1, 6.7) | 5.7 (4.6, 6.8) | 5.7 (5.1, 6.2) | 5.4 (4.1, 6.7) | 6.0 (5.2, 6.7) | 5.9 (4.8, 7.0) | 5.8 (5.3, 6.4) |
| Check-in 5 | N | 31 | 68 | 36 | 135 | 31 | 70 | 37 | 138 |
| | Mean (95%CIs) | 5.8 (4.3, 7.2) | 5.5 (4.6, 6.4) | 6.1 (4.8, 7.5) | 5.7 (5.1, 6.4) | 5.8 (4.3, 7.2) | 5.4 (4.5, 6.3) | 6.5 (5.0, 8.1) | 5.8 (5.1, 6.5) |
| Check-in 6 | N | 23 | 61 | 28 | 112 | 23 | 62 | 28 | 113 |
| | Mean (95%CIs) | 4.2 (2.9, 5.5) | 4.9 (3.9, 5.9) | 5.4 (3.7, 7.1) | 4.9 (4.2, 5.6) | 4.2 (2.9, 5.5) | 4.9 (4.0, 5.9) | 5.4 (3.7, 7.1) | 4.9 (4.2, 5.6) |
| Completion | N | 39 | 82 | 48 | 169 | 46 | 101 | 56 | 203 |
| | Mean (95%CIs) | 4.6 (3.5, 5.7) | 4.8 (4.0, 5.6) | 5.2 (4.0, 6.4) | 4.9 (4.3, 5.4) | 4.5 (3.5, 5.5) | 5.1 (4.3, 5.9) | 5.3 (4.2, 6.4) | 5.0 (4.5, 5.6) |
| Follow-up | N | 39 | 80 | 47 | 166 | 48 | 106 | 56 | 210 |
| | Mean (95%CIs) | 5.2 (3.9, 6.6) | 5.3 (4.2, 6.3) | 5.7 (4.1, 7.2) | 5.4 (4.7, 6.1) | 5.1 (4.0, 6.3) | 5.6 (4.7, 6.5) | 5.9 (4.5, 7.3) | 5.6 (4.9, 6.2) |
